# Prevalence and Sociodemographic Correlates of Depression and Thoughts of Self-Harm Among Adolescents in Five Low- and Middle-Income Countries: Evidence from the Demographic and Health Survey Program

**DOI:** 10.64898/2026.10.29.26366826

**Authors:** Lena Jäggi, Amit Aryal, Chimuka Muleya, Makamohelo Malimabe, Olga Cambaco, Günther Fink, Irene Falgas-Bague

## Abstract

**Background:** Adolescence is a critical period for the onset of depression and self-harm, yet comparable, nationally representative estimates from low- and middle-income countries (LMICs) remain scarce. Existing evidence relies heavily on school-based samples, excluding out-of-school adolescents. The recently available data from Demographic and Health Surveys (DHS) Mental Health Module offers an opportunity to address this gap.

**Methods and findings:** We analysed data from 16,269 adolescents aged 15–19 years across five DHS surveys: Bangladesh (2022), Lesotho (2023–24), Mozambique (2022–23), Nepal (2022), and Zambia (2024). Probable depression was defined as a PHQ-9 score ≥10, and thoughts of death or self-harm as any endorsement of PHQ-9 item 9. We estimated survey-weighted prevalence by country, sex, and age, examined symptom profiles, and used sex- and country-specific modified Poisson regression to identify sociodemographic correlates.

The country-normalised pooled prevalence was 3.9% (95% CI 3.4–4.5) for probable depression, ranging from 0.6% in Zambia to 6.5% in Mozambique, and 7.0% (95% CI 6.4–7.7) for thoughts of death or self-harm, ranging from 3.1% to 10.2%, respectively. Girls had higher prevalence than boys for both depression (4.9% vs 1.9%) and thoughts of death or self-harm (8.6% vs 3.9%), although sex differences varied across countries. No consistent age gradient emerged. Associations with household composition, marital and parental status, urban residence, education, and wealth were heterogeneous across settings.

Interpretation is limited by the lack of population-specific PHQ-9 validation and cross-country measurement invariance testing, the inability of item 9 to distinguish passive death wishes from suicidal ideation, the restriction to ever-married girls in Bangladesh, and the cross-sectional design.

**Conclusions:** Across five LMICs, PHQ-9-defined probable depression was relatively uncommon, while thoughts of death or self-harm affected approximately one in fourteen adolescents, particularly in girls. Substantial cross-country variation highlights the need for context-sensitive adolescent mental health surveillance and prevention, including out-of-school populations, supported by repeated household surveys and locally validated measures.

**Author summary:** **Why was this study done?**

- Depression and self-harm often begin during adolescence, and suicide is one of the leading causes of death among young people worldwide.
- In low- and middle-income countries, most of what we know comes from school-based or local studies that use different questionnaires. These studies miss adolescents who are not in school and are hard to compare across countries.
- The Demographic and Health Survey (DHS) Program recently added a mental health module to its national household surveys. This allows comparable estimates across countries for the first time.

**What did the researchers do and find?**

- We analysed responses from 16,269 adolescents aged 15–19 years in national household surveys from Bangladesh, Lesotho, Mozambique, Nepal and Zambia. All five surveys used the same questionnaire (PHQ-9) to assess depressive symptoms including suicide ideation and self-harm.
- About 4 in 100 adolescents had probable depression, ranging from under 1% in Zambia to 6.5% in Mozambique. Thoughts of death or self-harm were more common, affecting about 7 in 100 adolescents, and were more frequent than probable depression in every country.
- Girls reported more symptoms than boys in all countries, but the size of this gap varied widely between countries. Age, place of residence, education, wealth, marriage and parenthood showed no consistent pattern across countries.

**What do these findings mean?**

- Many adolescents in these countries have thoughts of death or self-harm, including some who do not meet the threshold for probable depression. Overall, girls report higher rates than boys. Screening based only on depression scores may miss them.
- Risk factors differ between countries, so prevention and services should be designed for each local context. They should also reach adolescents who are not in school.

## Introduction

Adolescence is a critical life-course period for mental health, during which many mental disorders first emerge and opportunities for prevention and early intervention may have lasting life-course benefits (1, 2). Depression is among the leading causes of illness and disability during adolescence, while suicide is the third leading cause of death among people aged 15–29 years globally (3). Recent reports from low- and middle-income countries (LMICs) have also documented a substantial, although highly heterogeneous, burden of self-harm and suicidal thoughts and behaviours, underscoring the need for improved surveillance, prevention, and equitable access to mental health care (4). In response, global calls for action increasingly frame youth mental health as a policy priority requiring coordinated prevention strategies, service reform, and attention to the broader social determinants of health (5).

The distribution of adolescent mental health problems remains poorly characterised in LMICs, where nationally representative population-based data are scarce. Much of the available literature is based on school-based or subnational studies using heterogeneous instruments, age ranges, and thresholds, making comparisons across settings difficult (6).Evidence generated during the COVID-19 pandemic also documented elevated depressive symptoms, often with higher levels among girls, reinforcing the need for recent population-based estimates from LMICs (7–10). Moreover, most cross-national studies focus on the total adult population, with limited attention to how depressive symptoms vary across the adolescent years, which specific symptoms contribute most to the overall burden, or how symptom burden differs across key sociodemographic groups. At the same time, adolescents are experiencing rapid demographic, social, and technological changes that may influence mental health differently across settings. Comparable population-based surveillance data are therefore needed to describe geographic and socioeconomic variation, identify populations at greatest risk, and inform prevention and intervention strategies (11).

The introduction of the Mental Health Module into the Demographic and Health Survey (DHS) program represents an important opportunity for the standardized population-based adolescent mental health surveillance in LMICs. The DHS Program has conducted nationally representative household surveys in more than 70 countries since the 1980s and is a key source of maternal and child health indicators (12). Unlike school-based surveys, DHS household samples can include adolescents both in and out of school. In a field where global narratives are often informed by heterogeneous measures, non-comparable studies, and evolving reporting practices, comparable descriptive benchmarks strengthen the evidence base for adolescent mental health surveillance in LMICs, directly addressing long-standing concerns regarding the measurement and interpretation of global mental health trends (13). However, the current mental health module captures self-reported symptoms of depression and suicidal ideation rather than clinical diagnoses and cross-country interpretation must account for the limitations inherent to symptom-based measures, including potential cultural variation in symptom expression and contextual adaptation across settings.

Previous studies have established a substantial but highly heterogeneous burden of depression and suicidal thoughts and behaviours among adolescents in LMICs. A systematic review of 37 studies involving 97,616 adolescents in sub-Saharan Africa reported median prevalences of 26.9% for depression and 20.8% for suicidal ideation, but also highlighted substantial variation in sampling approaches, settings and measurement instruments (14). Similarly, Uddin et al. (2019) (15), using nationally representative school-based surveys of 229,129 adolescents aged 13–17 years across 59 LMICs, estimated a 16.9% prevalence of suicidal ideation and found higher prevalence among girls and older adolescents; however, the study was restricted to school-going adolescents and did not concurrently characterise depressive symptom profiles. More recently, the National Adolescent Mental Health Surveys used nationally representative household samples in Kenya, Indonesia and Viet Nam to assess DSM-5 mental disorders together with self-harm and suicidal behaviours among adolescents aged 10–17 years, demonstrating marked between-country variation but not addressing late adolescence or PHQ-9 symptom-level variation (16)). A further cross-country analysis of 12 LMICs examined depression, self-harm and suicidal ideation among internet-using adolescents aged 12–17 years and found considerable heterogeneity in both prevalence and sociodemographic patterns, while also failing to establish measurement invariance across countries (17). Collectively, these studies demonstrate the importance of adolescent mental health in LMICs but leave limited comparable, recent household-based evidence focused specifically on late adolescence, including adolescents outside school, using a common instrument to examine depression, individual symptom patterns, thoughts of death or self-harm, and sociodemographic variation across countries, the gaps that the present DHS analysis seeks to address.

This study uses the first five DHS surveys to include the Mental Health Module with nationally representative data from Bangladesh, Lesotho, Mozambique, Nepal, and Zambia to provide standardized population-based estimates of probable depression and thoughts of death or self-harm among adolescents aged 15–19 years. We also identify key sociodemographic factors associated with adolescent mental health problems to inform more targeted surveillance, prevention, and mental health services.

## Methods

### Study design and data sources

We conducted a multi-country, cross-sectional secondary data analysis of nationally representative data collected within the following surveys: Bangladesh DHS 2022, Lesotho DHS 2023–24, Mozambique DHS 2022–23, Nepal DHS 2022, Zambia DHS 2024.

### Study population

The analytic sample comprised all respondents aged 15–19 years who were eligible for the mental health module under each country’s survey design and had valid data for mental health outcomes. In Mozambique, Zambia, Lesotho, and Nepal, the module was administered to both female and male respondents, and analyses were therefore stratified by sex. In Bangladesh, the module was administered only to ever-married female respondents; analyses for Bangladesh were consequently restricted to this population and are not directly representative of all adolescent girls aged 15–19 years.

To maintain a consistent denominator across outcomes, the final analytic sample was restricted to adolescents with both a valid PHQ-9 total score and a valid response to PHQ-9 item 9. Respondents outside the eligible age range, those not selected for the mental health module, and those without valid data for either outcome were excluded.

### Measures

#### Outcomes

We examined two mental health outcomes included in the mental health module separately: probable depression and thoughts of death or self-harm. They were assessed using the PHQ-9, a widely used screening measure of depressive symptoms (18). PHQ-9 consists of nine specific questions about depressive symptoms experienced in the past two weeks including anhedonia, depressed mood, sleep disturbances, fatigue, appetite changes, feelings of worthlessness, concentration difficulties, psychomotor changes, and thoughts of self-harm. Responses on the 9-item questionnaire were coded on the standard four-point frequency scale: 0 (“not at all”), 1 (“several days”), 2 (more than half the days”, and 3 (“nearly every day”). Responses recorded as “refused” or “don’t know” were treated as missing rather than recoded as absence of symptoms. The number of valid PHQ-9 items was first calculated for each respondent. A total score was derived for adolescents with valid responses to at least eight of the nine items. For respondents with exactly one missing item, the sum of the available items was prorated to the nine-item scale. Respondents with fewer than eight valid items were excluded from analyses. Probable depression was defined as a prorated PHQ-9 total score of 10 or higher, consistent with previous literature (18). Additionally, overall PHQ-9 symptom severity and single item responses were summarized descriptively where relevant.

Thoughts of death or self-harm were assessed using PHQ-9 item 9, which asks about thoughts that the respondent would be better off dead or of hurting themselves during the previous two weeks. Item 9 responses coded as “refused” or “don’t know” were treated as missing. The primary screen -positive definition was any valid response greater than zero, corresponding to endorsement of “several days” or more during the preceding two weeks.

Country questionnaires of PHQ-9 were adapted from DHS model instruments and administered in locally translated versions: Bangla in Bangladesh; Nepali, Maithili and Bhojpuri in Nepal; Portuguese in Mozambique; English and Sesotho in Lesotho; and English, Bemba, Kaonde, Lozi, Lunda, Luvale, Nyanja and Tonga in Zambia (19–30). The extent of documented PHQ-9 adaptation and validation differed across surveys. In Nepal, the DHS team worked with the Transcultural Psychosocial Organization Nepal to incorporate culturally meaningful terminology and used a Water Glass Pictorial Scale to support interpretation of response frequencies (21, 26). Related studies have criterion-validated culturally adapted Nepali PHQ versions among adults and adolescents (23, 24). Country-specific PHQ-9 validation studies are also available from Bangladesh and Mozambique (25, 26), although available survey documentation does not confirm that the DHS used the exact versions evaluated in those studies. For Lesotho and Zambia, we found documentation of local translation and survey adaptation but no published criterion validation of the exact PHQ-9 versions used by DHS (22, 29).

#### Stratification variables

All estimates were stratified by sex (as recorded in DHS) where the mental health module was available for both sexes. Age gradients were assessed using single-year age (15–19 years).

#### Covariates

The primary adjusted models included place of residence, educational attainment, household wealth, and household composition.

Place of residence was classified as urban or rural using DHS variable v025. Educational attainment was derived from v106 and grouped as no or basic education versus secondary education or higher. Household wealth was based on the DHS household wealth index (v190), and grouped into poorest or poorer, middle, and richer or richest categories. Household composition was derived using a prespecified algorithm based primarily on the respondent’s relationship to the household head. Adolescents were classified as living with parents or grandparents; living with in-laws or as the household head or spouse; or living in another household arrangement. Marital status was derived from v501 and classified as never married, currently married or in union, separated, divorced, or widowed. Parenthood was defined as ever having had a live birth or being currently pregnant, derived using variables v201 and v213.

### Statistical analysis

All analyses accounted for the DHS complex survey design, incorporating sampling weights, clustering at the PSU level, and stratification. The primary estimand was country-specific prevalence and associations, interpreted as nationally representative within each survey.

For cross-country descriptive comparisons in prevalence tables, we appended surveys and constructed a country-normalised pooled dataset by rescaling each country’s survey weights to sum to 1; this pooled column is presented as a comparative country-average benchmark.

We estimated weighted prevalence and design-based 95% CIs for (1) probable depression (PHQ-9 ≥10) and (2) thoughts of death or self-harm (PHQ-9 item 9 ≥1), overall and by sex where available, and report unweighted denominators. Age gradients were assessed using single-year age (15–19 years) within countries (sex stratified where available) and displayed as country-specific age curves; tests for age heterogeneity are reported in the appendix, table S2.

To characterize symptom profiles, we estimated the weighted percentage endorsing each PHQ-9 item at ≥1 (“several days” or more) by country and sex, and repeated these analyses among adolescents with PHQ-9 ≥10. Results are included in figure 2; corresponding numeric estimates are provided in the appendix table S3.

For sociodemographic correlates, we fit country-specific modified Poisson regression models with robust variance to estimate adjusted prevalence ratios (aPRs) for each outcome, stratified by sex where available. Covariates were prespecified and entered as domains (place of residence; socioeconomic position; household composition). Regression coefficients were not pooled; country-specific aPRs are presented side by side to assess consistency and heterogeneity. Supplementary analyses among girls additionally included marital or union status and parenthood. Girls who were separated, divorced, or widowed were excluded from these supplementary models due to extremely low cell counts.

Because PHQ-9 cut-points reflect a screening decision rule rather than a fixed diagnostic boundary, we conducted a prespecified sensitivity analysis for depression prevalence using an alternative cut-point (PHQ-9 ≥8). This choice was motivated by adolescent-focused evidence from Mozambique and other sub-Saharan African countries(31) and by precedent in an analysis of the 2022–23 Mozambique DHS adolescent mental health module that applied a cutoff of 8, citing acceptable sensitivity and specificity in Mozambican adolescents. Estimates from this sensitivity analysis are presented in the appendix to assess robustness of descriptive conclusions to the choice of threshold (figure S5) (32, 33).

## Results

### Adolescent characteristics

Table 1 summarizes the demographic characteristics of the pooled study population and each participating country. The pooled sample included 16,269 adolescents aged 15–19 years, with females comprising 65.9% of the sample. Overall, 56.3% of participants lived in rural areas and 61.7% had attained secondary education, although substantial variation was observed across countries. Nepal had the highest proportion of urban adolescents (68.1%), whereas Bangladesh was predominantly rural (76.3%). Educational attainment was lowest in Mozambique, where 12.5% of adolescents had no formal education and 44.4% had only primary education, compared with Bangladesh and Lesotho, where approximately three-quarters had completed some secondary education. Most adolescents lived with their parents or grandparents, ranging from 56.8% in Mozambique to 77.8% in Nepal, whereas a quarter of the adolescents were head or spouse of the household in Bangladesh. The proportion of adolescents who were currently married varied widely, from 5.7% in Lesotho to 98.1% in Bangladesh, reflecting the survey design in Bangladesh, while adolescent parenthood ranged from 5.9% in Nepal to 53.0% in Bangladesh.

**Table 1.**
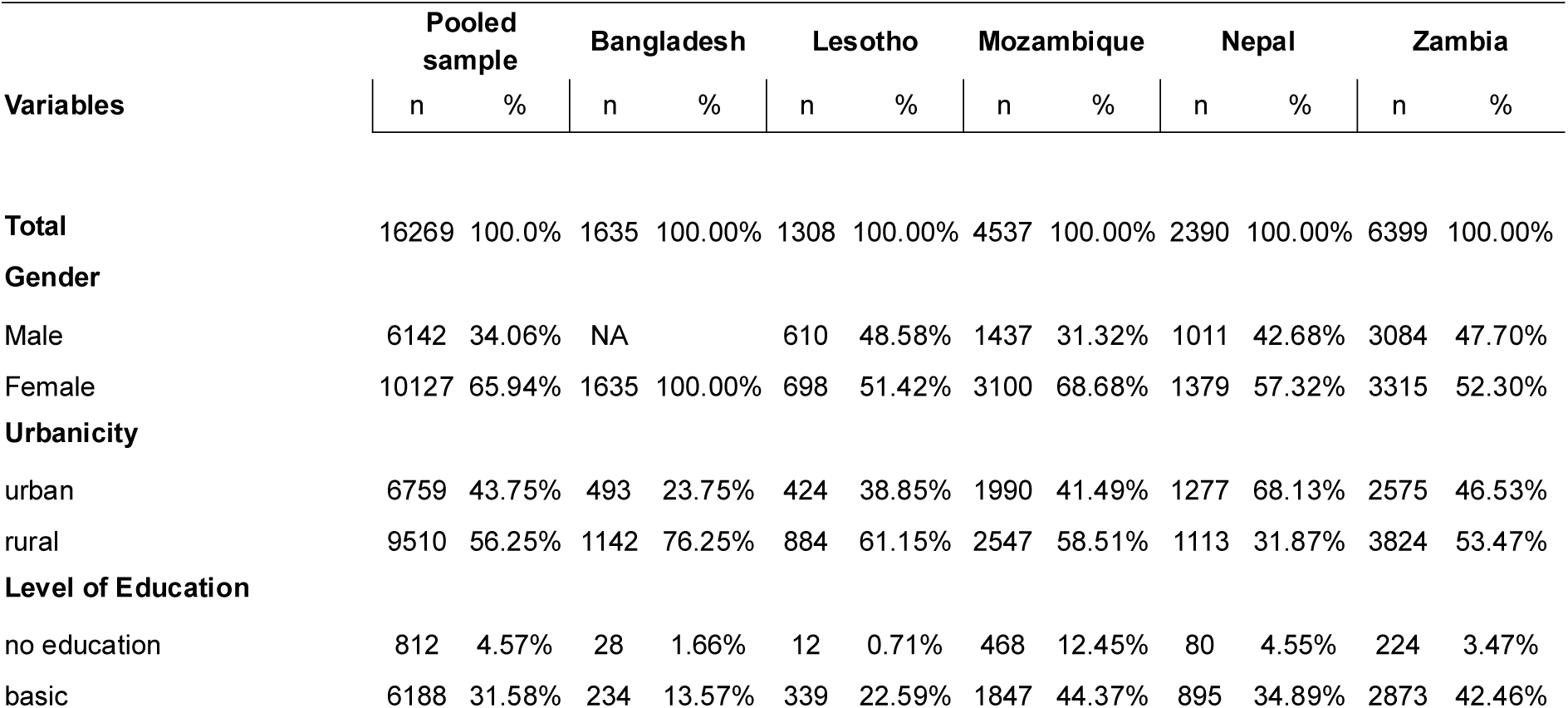
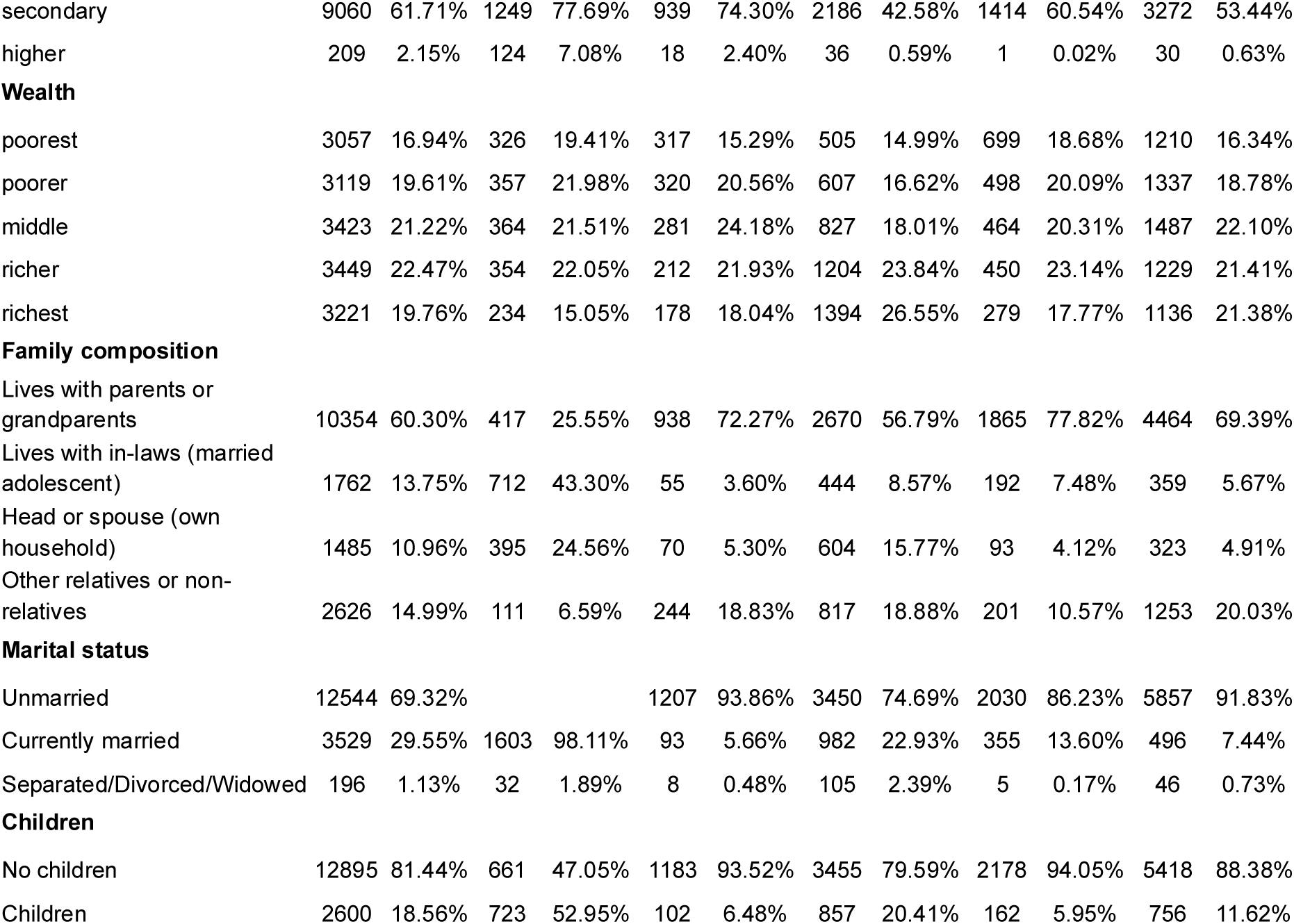
Demographic characteristics of all Adolescents included in the DHS survey.

|  | **Pooled sample** |  | **Bangladesh** |  | **Lesotho** |  | **Mozambique** |  | **Nepal** |  | **Zambia** |  |
|---|---|---|---|---|---|---|---|---|---|---|---|---|
| **Variables** | n | % | n | % | n | % | n | % | n | % | n | % |
| **Total** | 16269 | 100.0% | 1635 | 100.00% | 1308 | 100.00% | 4537 | 100.00% | 2390 | 100.00% | 6399 | 100.00% |
| **Gender** |  |  |  |  |  |  |  |  |  |  |  |  |
| Male | 6142 | 34.06% | NA |  | 610 | 48.58% | 1437 | 31.32% | 1011 | 42.68% | 3084 | 47.70% |
| Female | 10127 | 65.94% | 1635 | 100.00% | 698 | 51.42% | 3100 | 68.68% | 1379 | 57.32% | 3315 | 52.30% |
| **Urbanicity** |  |  |  |  |  |  |  |  |  |  |  |  |
| urban | 6759 | 43.75% | 493 | 23.75% | 424 | 38.85% | 1990 | 41.49% | 1277 | 68.13% | 2575 | 46.53% |
| rural | 9510 | 56.25% | 1142 | 76.25% | 884 | 61.15% | 2547 | 58.51% | 1113 | 31.87% | 3824 | 53.47% |
| **Level of Education** |  |  |  |  |  |  |  |  |  |  |  |  |
| no education | 812 | 4.57% | 28 | 1.66% | 12 | 0.71% | 468 | 12.45% | 80 | 4.55% | 224 | 3.47% |
| basic | 6188 | 31.58% | 234 | 13.57% | 339 | 22.59% | 1847 | 44.37% | 895 | 34.89% | 2873 | 42.46% |
| secondary | 9060 | 61.71% | 1249 | 77.69% | 939 | 74.30% | 2186 | 42.58% | 1414 | 60.54% | 3272 | 53.44% |
| higher | 209 | 2.15% | 124 | 7.08% | 18 | 2.40% | 36 | 0.59% | 1 | 0.02% | 30 | 0.63% |
| **Wealth** |  |  |  |  |  |  |  |  |  |  |  |  |
| poorest | 3057 | 16.94% | 326 | 19.41% | 317 | 15.29% | 505 | 14.99% | 699 | 18.68% | 1210 | 16.34% |
| poorer | 3119 | 19.61% | 357 | 21.98% | 320 | 20.56% | 607 | 16.62% | 498 | 20.09% | 1337 | 18.78% |
| middle | 3423 | 21.22% | 364 | 21.51% | 281 | 24.18% | 827 | 18.01% | 464 | 20.31% | 1487 | 22.10% |
| richer | 3449 | 22.47% | 354 | 22.05% | 212 | 21.93% | 1204 | 23.84% | 450 | 23.14% | 1229 | 21.41% |
| richest | 3221 | 19.76% | 234 | 15.05% | 178 | 18.04% | 1394 | 26.55% | 279 | 17.77% | 1136 | 21.38% |
| **Family composition** |  |  |  |  |  |  |  |  |  |  |  |  |
| Lives with parents or grandparents | 10354 | 60.30% | 417 | 25.55% | 938 | 72.27% | 2670 | 56.79% | 1865 | 77.82% | 4464 | 69.39% |
| Lives with in-laws (married adolescent) | 1762 | 13.75% | 712 | 43.30% | 55 | 3.60% | 444 | 8.57% | 192 | 7.48% | 359 | 5.67% |
| Head or spouse (own household) | 1485 | 10.96% | 395 | 24.56% | 70 | 5.30% | 604 | 15.77% | 93 | 4.12% | 323 | 4.91% |
| Other relatives or non-relatives | 2626 | 14.99% | 111 | 6.59% | 244 | 18.83% | 817 | 18.88% | 201 | 10.57% | 1253 | 20.03% |
| **Marital status** |  |  |  |  |  |  |  |  |  |  |  |  |
| Unmarried | 12544 | 69.32% |  |  | 1207 | 93.86% | 3450 | 74.69% | 2030 | 86.23% | 5857 | 91.83% |
| Currently married | 3529 | 29.55% | 1603 | 98.11% | 93 | 5.66% | 982 | 22.93% | 355 | 13.60% | 496 | 7.44% |
| Separated/Divorced/Widowed | 196 | 1.13% | 32 | 1.89% | 8 | 0.48% | 105 | 2.39% | 5 | 0.17% | 46 | 0.73% |
| **Children** |  |  |  |  |  |  |  |  |  |  |  |  |
| No children | 12895 | 81.44% | 661 | 47.05% | 1183 | 93.52% | 3455 | 79.59% | 2178 | 94.05% | 5418 | 88.38% |
| Children | 2600 | 18.56% | 723 | 52.95% | 102 | 6.48% | 857 | 20.41% | 162 | 5.95% | 756 | 11.62% |

### Probable depression by demographic characteristics

Table 2 presents the weighted prevalence of probable depression (PHQ-9 ≥10) by demographic characteristics overall and by country. In the pooled country-normalized sample, the prevalence of probable depression was 3.9% (95% CI 3.4–4.5), ranging from 0.6% in Zambia to 6.5% in Mozambique, with intermediate estimates in Lesotho (5.6%), Bangladesh (3.7%), and Nepal (3.3%).

**Table 2.**
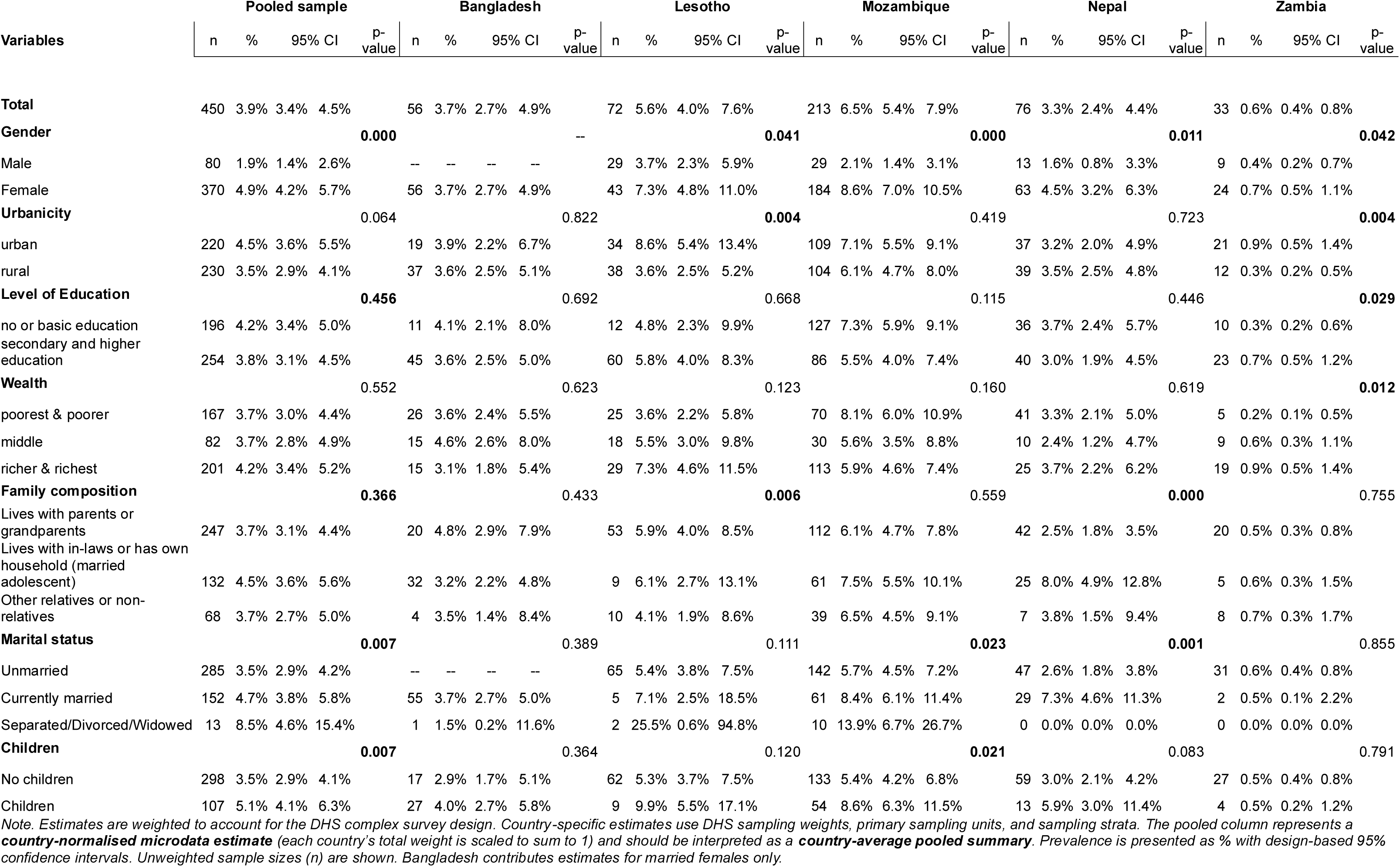
Weighted prevalence of probable depression (PHQ-9≥10) among adolescents by demographic characteristics, pooled and by country.

|  | Pooled sample |  |  |  |  | Bangladesh |  |  |  |  | Lesotho |  |  |  |  | Mozambique |  |  |  |  | Nepal |  |  |  |  | Zambia |  |  |  |  |
|---|---|---|---|---|---|---|---|---|---|---|---|---|---|---|---|---|---|---|---|---|---|---|---|---|---|---|---|---|---|---|
| Variables | n | % | 95% CI |  | p-value | n | % | 95% CI |  | p-value | n | % | 95% CI |  | p-value | n | % | 95% CI |  | p-value | n | % | 95% CI |  | p-value | n | % | 95% CI |  | p-value |
| **Total** | 450 | 3.9% | 3.4% | 4.5% |  | 56 | 3.7% | 2.7% | 4.9% |  | 72 | 5.6% | 4.0% | 7.6% |  | 213 | 6.5% | 5.4% | 7.9% |  | 76 | 3.3% | 2.4% | 4.4% |  | 33 | 0.6% | 0.4% | 0.8% |  |
| **Gender** |  |  |  |  | **0.000** |  |  |  |  | -- |  |  |  |  | **0.041** |  |  |  |  | **0.000** |  |  |  |  | **0.011** |  |  |  |  | **0.042** |
| Male | 80 | 1.9% | 1.4% | 2.6% |  | -- | -- | -- | -- |  | 29 | 3.7% | 2.3% | 5.9% |  | 29 | 2.1% | 1.4% | 3.1% |  | 13 | 1.6% | 0.8% | 3.3% |  | 9 | 0.4% | 0.2% | 0.7% |  |
| Female | 370 | 4.9% | 4.2% | 5.7% |  | 56 | 3.7% | 2.7% | 4.9% |  | 43 | 7.3% | 4.8% | 11.0% |  | 184 | 8.6% | 7.0% | 10.5% |  | 63 | 4.5% | 3.2% | 6.3% |  | 24 | 0.7% | 0.5% | 1.1% |  |
| **Urbanicity** |  |  |  |  | 0.064 |  |  |  |  | 0.822 |  |  |  |  | **0.004** |  |  |  |  | 0.419 |  |  |  |  | 0.723 |  |  |  |  | **0.004** |
| urban | 220 | 4.5% | 3.6% | 5.5% |  | 19 | 3.9% | 2.2% | 6.7% |  | 34 | 8.6% | 5.4% | 13.4% |  | 109 | 7.1% | 5.5% | 9.1% |  | 37 | 3.2% | 2.0% | 4.9% |  | 21 | 0.9% | 0.5% | 1.4% |  |
| rural | 230 | 3.5% | 2.9% | 4.1% |  | 37 | 3.6% | 2.5% | 5.1% |  | 38 | 3.6% | 2.5% | 5.2% |  | 104 | 6.1% | 4.7% | 8.0% |  | 39 | 3.5% | 2.5% | 4.8% |  | 12 | 0.3% | 0.2% | 0.5% |  |
| **Level of Education** |  |  |  |  | **0.456** |  |  |  |  | 0.692 |  |  |  |  | 0.668 |  |  |  |  | 0.115 |  |  |  |  | 0.446 |  |  |  |  | **0.029** |
| no or basic education | 196 | 4.2% | 3.4% | 5.0% |  | 11 | 4.1% | 2.1% | 8.0% |  | 12 | 4.8% | 2.3% | 9.9% |  | 127 | 7.3% | 5.9% | 9.1% |  | 36 | 3.7% | 2.4% | 5.7% |  | 10 | 0.3% | 0.2% | 0.6% |  |
| secondary and higher education | 254 | 3.8% | 3.1% | 4.5% |  | 45 | 3.6% | 2.5% | 5.0% |  | 60 | 5.8% | 4.0% | 8.3% |  | 86 | 5.5% | 4.0% | 7.4% |  | 40 | 3.0% | 1.9% | 4.5% |  | 23 | 0.7% | 0.5% | 1.2% |  |
| **Wealth** |  |  |  |  | 0.552 |  |  |  |  | 0.623 |  |  |  |  | 0.123 |  |  |  |  | 0.160 |  |  |  |  | 0.619 |  |  |  |  | **0.012** |
| poorest & poorer | 167 | 3.7% | 3.0% | 4.4% |  | 26 | 3.6% | 2.4% | 5.5% |  | 25 | 3.6% | 2.2% | 5.8% |  | 70 | 8.1% | 6.0% | 10.9% |  | 41 | 3.3% | 2.1% | 5.0% |  | 5 | 0.2% | 0.1% | 0.5% |  |
| middle | 82 | 3.7% | 2.8% | 4.9% |  | 15 | 4.6% | 2.6% | 8.0% |  | 18 | 5.5% | 3.0% | 9.8% |  | 30 | 5.6% | 3.5% | 8.8% |  | 10 | 2.4% | 1.2% | 4.7% |  | 9 | 0.6% | 0.3% | 1.1% |  |
| richer & richest | 201 | 4.2% | 3.4% | 5.2% |  | 15 | 3.1% | 1.8% | 5.4% |  | 29 | 7.3% | 4.6% | 11.5% |  | 113 | 5.9% | 4.6% | 7.4% |  | 25 | 3.7% | 2.2% | 6.2% |  | 19 | 0.9% | 0.5% | 1.4% |  |
| **Family composition** |  |  |  |  | **0.366** |  |  |  |  | 0.433 |  |  |  |  | **0.006** |  |  |  |  | 0.559 |  |  |  |  | **0.000** |  |  |  |  | 0.755 |
| Lives with parents or grandparents | 247 | 3.7% | 3.1% | 4.4% |  | 20 | 4.8% | 2.9% | 7.9% |  | 53 | 5.9% | 4.0% | 8.5% |  | 112 | 6.1% | 4.7% | 7.8% |  | 42 | 2.5% | 1.8% | 3.5% |  | 20 | 0.5% | 0.3% | 0.8% |  |
| Lives with in-laws or has own household (married adolescent) | 132 | 4.5% | 3.6% | 5.6% |  | 32 | 3.2% | 2.2% | 4.8% |  | 9 | 6.1% | 2.7% | 13.1% |  | 61 | 7.5% | 5.5% | 10.1% |  | 25 | 8.0% | 4.9% | 12.8% |  | 5 | 0.6% | 0.3% | 1.5% |  |
| Other relatives or non-relatives | 68 | 3.7% | 2.7% | 5.0% |  | 4 | 3.5% | 1.4% | 8.4% |  | 10 | 4.1% | 1.9% | 8.6% |  | 39 | 6.5% | 4.5% | 9.1% |  | 7 | 3.8% | 1.5% | 9.4% |  | 8 | 0.7% | 0.3% | 1.7% |  |
| **Marital status** |  |  |  |  | **0.007** |  |  |  |  | 0.389 |  |  |  |  | 0.111 |  |  |  |  | **0.023** |  |  |  |  | **0.001** |  |  |  |  | 0.855 |
| Unmarried | 285 | 3.5% | 2.9% | 4.2% |  | -- | -- | -- | -- |  | 65 | 5.4% | 3.8% | 7.5% |  | 142 | 5.7% | 4.5% | 7.2% |  | 47 | 2.6% | 1.8% | 3.8% |  | 31 | 0.6% | 0.4% | 0.8% |  |
| Currently married | 152 | 4.7% | 3.8% | 5.8% |  | 55 | 3.7% | 2.7% | 5.0% |  | 5 | 7.1% | 2.5% | 18.5% |  | 61 | 8.4% | 6.1% | 11.4% |  | 29 | 7.3% | 4.6% | 11.3% |  | 2 | 0.5% | 0.1% | 2.2% |  |
| Separated/Divorced/Widowed | 13 | 8.5% | 4.6% | 15.4% |  | 1 | 1.5% | 0.2% | 11.6% |  | 2 | 25.5% | 0.6% | 94.8% |  | 10 | 13.9% | 6.7% | 26.7% |  | 0 | 0.0% | 0.0% | 0.0% |  | 0 | 0.0% | 0.0% | 0.0% |  |
| **Children** |  |  |  |  | **0.007** |  |  |  |  | 0.364 |  |  |  |  | 0.120 |  |  |  |  | **0.021** |  |  |  |  | 0.083 |  |  |  |  | 0.791 |
| No children | 298 | 3.5% | 2.9% | 4.1% |  | 17 | 2.9% | 1.7% | 5.1% |  | 62 | 5.3% | 3.7% | 7.5% |  | 133 | 5.4% | 4.2% | 6.8% |  | 59 | 3.0% | 2.1% | 4.2% |  | 27 | 0.5% | 0.4% | 0.8% |  |
| Children | 107 | 5.1% | 4.1% | 6.3% |  | 27 | 4.0% | 2.7% | 5.8% |  | 9 | 9.9% | 5.5% | 17.1% |  | 54 | 8.6% | 6.3% | 11.5% |  | 13 | 5.9% | 3.0% | 11.4% |  | 4 | 0.5% | 0.2% | 1.2% |  |

*Note. Estimates are weighted to account for the DHS complex survey design. Country-specific estimates use DHS sampling weights, primary sampling units, and sampling strata. The pooled column represents a* ***country-normalised microdata estimate*** *(each country’s total weight is scaled to sum to 1) and should be interpreted as a* ***country-average pooled summary****. Prevalence is presented as % with design-based 95% confidence intervals. Unweighted sample sizes (n) are shown. Bangladesh contributes estimates for married females only.*

Across the pooled sample, females had a significantly higher prevalence of probable depression than males (4.9% vs. 1.9%; p<0.001). This pattern was consistently observed in Lesotho, Mozambique, Nepal, and Zambia, although statistical significance varied across countries. Urbanicity, educational attainment, and household wealth were not associated with probable depression in the pooled analysis, although significant country-specific differences were observed in Lesotho and Zambia.

In the pooled sample, prevalence differed by marital status and parenthood, with higher prevalence among adolescents who were currently married (4.7%) or separated/divorced/widowed (8.5%) than among unmarried adolescents (3.5%; p=0.007). Similarly, adolescents with children had a higher prevalence of probable depression than those without children (5.1% vs. 3.5%; p=0.007). Associations with family composition, marital status, and parenthood varied across countries, with the strongest differences observed in Nepal and Mozambique.

### Thoughts of death or self-harm by demographic characteristics

Table 3 presents the weighted prevalence of thoughts of death or self-harm (PHQ-9 item 9 ≥1) by demographic characteristics. In the pooled country-normalized sample, the prevalence was 7.0% (95% CI 6.4–7.7), ranging from 3.1% in Zambia to 10.2% in Mozambique, with similarly high estimates in Lesotho (9.9%) and Bangladesh (7.9%), and a lower prevalence in Nepal (4.0%).

**Table 3.**
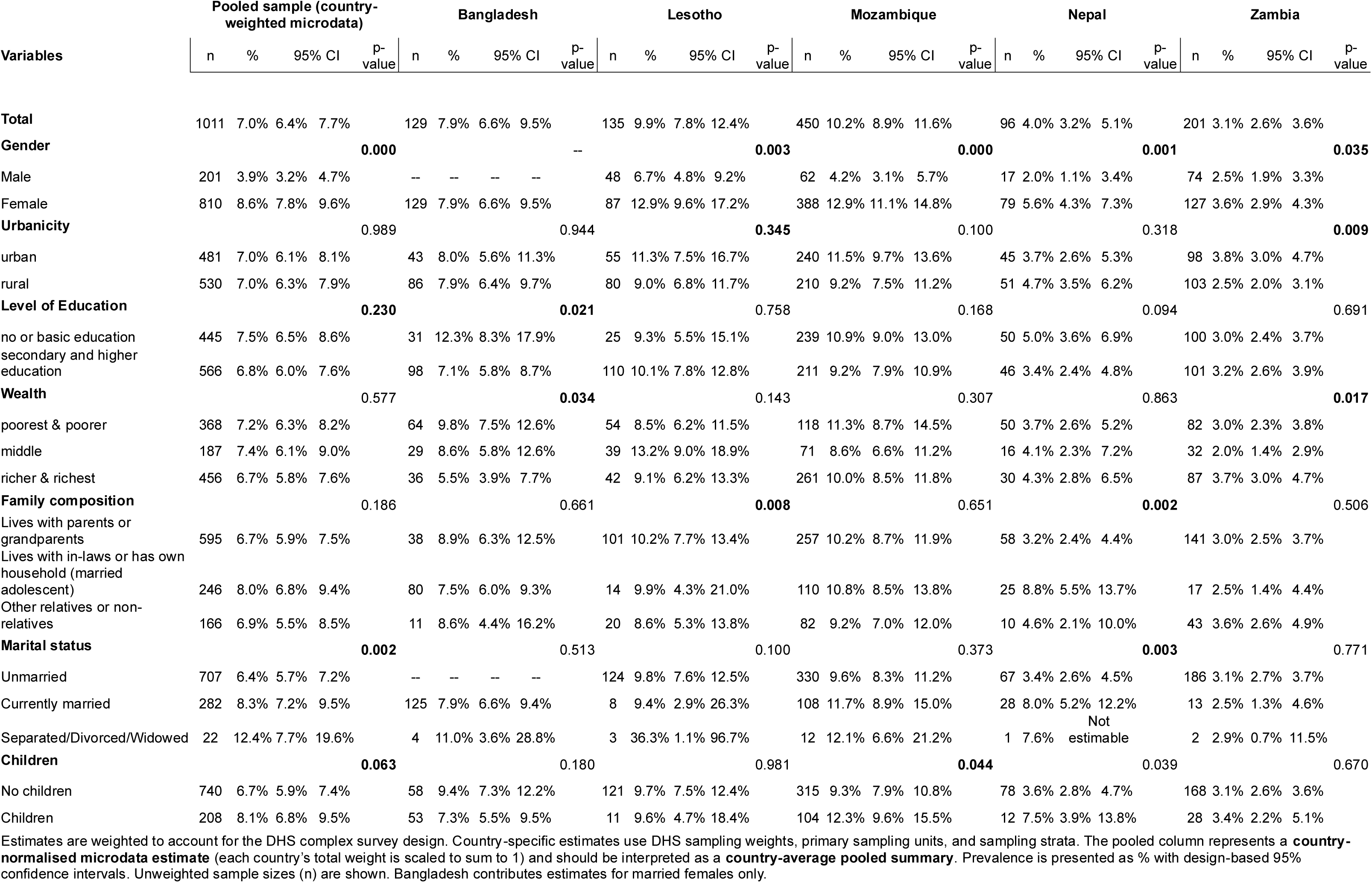
Prevalence of thoughts of death/self-harm (PHQ-9 item 9 ≥1) among adolescents by demographic characteristics, pooled and by country.

|  | Pooled sample (country-weighted microdata) |  |  |  |  | Bangladesh |  |  |  |  | Lesotho |  |  |  |  | Mozambique |  |  |  |  | Nepal |  |  |  |  | Zambia |  |  |  |  |
|---|---|---|---|---|---|---|---|---|---|---|---|---|---|---|---|---|---|---|---|---|---|---|---|---|---|---|---|---|---|---|
| **Variables** | n | % | 95% CI |  | p-value | n | % | 95% CI |  | p-value | n | % | 95% CI |  | p-value | n | % | 95% CI |  | p-value | n | % | 95% CI |  | p-value | n | % | 95% CI |  | p-value |
| **Total** | 1011 | 7.0% | 6.4% | 7.7% |  | 129 | 7.9% | 6.6% | 9.5% |  | 135 | 9.9% | 7.8% | 12.4% |  | 450 | 10.2% | 8.9% | 11.6% |  | 96 | 4.0% | 3.2% | 5.1% |  | 201 | 3.1% | 2.6% | 3.6% |  |
| **Gender** |  |  |  |  | **0.000** |  |  |  |  | -- |  |  |  |  | **0.003** |  |  |  |  | **0.000** |  |  |  |  | **0.001** |  |  |  |  | **0.035** |
| Male | 201 | 3.9% | 3.2% | 4.7% |  | -- | -- | -- | -- |  | 48 | 6.7% | 4.8% | 9.2% |  | 62 | 4.2% | 3.1% | 5.7% |  | 17 | 2.0% | 1.1% | 3.4% |  | 74 | 2.5% | 1.9% | 3.3% |  |
| Female | 810 | 8.6% | 7.8% | 9.6% |  | 129 | 7.9% | 6.6% | 9.5% |  | 87 | 12.9% | 9.6% | 17.2% |  | 388 | 12.9% | 11.1% | 14.8% |  | 79 | 5.6% | 4.3% | 7.3% |  | 127 | 3.6% | 2.9% | 4.3% |  |
| **Urbanicity** |  |  |  |  | 0.989 |  |  |  |  | 0.944 |  |  |  |  | **0.345** |  |  |  |  | 0.100 |  |  |  |  | 0.318 |  |  |  |  | **0.009** |
| urban | 481 | 7.0% | 6.1% | 8.1% |  | 43 | 8.0% | 5.6% | 11.3% |  | 55 | 11.3% | 7.5% | 16.7% |  | 240 | 11.5% | 9.7% | 13.6% |  | 45 | 3.7% | 2.6% | 5.3% |  | 98 | 3.8% | 3.0% | 4.7% |  |
| rural | 530 | 7.0% | 6.3% | 7.9% |  | 86 | 7.9% | 6.4% | 9.7% |  | 80 | 9.0% | 6.8% | 11.7% |  | 210 | 9.2% | 7.5% | 11.2% |  | 51 | 4.7% | 3.5% | 6.2% |  | 103 | 2.5% | 2.0% | 3.1% |  |
| **Level of Education** |  |  |  |  | **0.230** |  |  |  |  | **0.021** |  |  |  |  | 0.758 |  |  |  |  | 0.168 |  |  |  |  | 0.094 |  |  |  |  | 0.691 |
| no or basic education | 445 | 7.5% | 6.5% | 8.6% |  | 31 | 12.3% | 8.3% | 17.9% |  | 25 | 9.3% | 5.5% | 15.1% |  | 239 | 10.9% | 9.0% | 13.0% |  | 50 | 5.0% | 3.6% | 6.9% |  | 100 | 3.0% | 2.4% | 3.7% |  |
| secondary and higher education | 566 | 6.8% | 6.0% | 7.6% |  | 98 | 7.1% | 5.8% | 8.7% |  | 110 | 10.1% | 7.8% | 12.8% |  | 211 | 9.2% | 7.9% | 10.9% |  | 46 | 3.4% | 2.4% | 4.8% |  | 101 | 3.2% | 2.6% | 3.9% |  |
| **Wealth** |  |  |  |  | 0.577 |  |  |  |  | **0.034** |  |  |  |  | 0.143 |  |  |  |  | 0.307 |  |  |  |  | 0.863 |  |  |  |  | **0.017** |
| poorest & poorer | 368 | 7.2% | 6.3% | 8.2% |  | 64 | 9.8% | 7.5% | 12.6% |  | 54 | 8.5% | 6.2% | 11.5% |  | 118 | 11.3% | 8.7% | 14.5% |  | 50 | 3.7% | 2.6% | 5.2% |  | 82 | 3.0% | 2.3% | 3.8% |  |
| middle | 187 | 7.4% | 6.1% | 9.0% |  | 29 | 8.6% | 5.8% | 12.6% |  | 39 | 13.2% | 9.0% | 18.9% |  | 71 | 8.6% | 6.6% | 11.2% |  | 16 | 4.1% | 2.3% | 7.2% |  | 32 | 2.0% | 1.4% | 2.9% |  |
| richer & richest | 456 | 6.7% | 5.8% | 7.6% |  | 36 | 5.5% | 3.9% | 7.7% |  | 42 | 9.1% | 6.2% | 13.3% |  | 261 | 10.0% | 8.5% | 11.8% |  | 30 | 4.3% | 2.8% | 6.5% |  | 87 | 3.7% | 3.0% | 4.7% |  |
| **Family composition** |  |  |  |  | 0.186 |  |  |  |  | 0.661 |  |  |  |  | **0.008** |  |  |  |  | 0.651 |  |  |  |  | **0.002** |  |  |  |  | 0.506 |
| Lives with parents or grandparents | 595 | 6.7% | 5.9% | 7.5% |  | 38 | 8.9% | 6.3% | 12.5% |  | 101 | 10.2% | 7.7% | 13.4% |  | 257 | 10.2% | 8.7% | 11.9% |  | 58 | 3.2% | 2.4% | 4.4% |  | 141 | 3.0% | 2.5% | 3.7% |  |
| Lives with in-laws or has own household (married adolescent) | 246 | 8.0% | 6.8% | 9.4% |  | 80 | 7.5% | 6.0% | 9.3% |  | 14 | 9.9% | 4.3% | 21.0% |  | 110 | 10.8% | 8.5% | 13.8% |  | 25 | 8.8% | 5.5% | 13.7% |  | 17 | 2.5% | 1.4% | 4.4% |  |
| Other relatives or non-relatives | 166 | 6.9% | 5.5% | 8.5% |  | 11 | 8.6% | 4.4% | 16.2% |  | 20 | 8.6% | 5.3% | 13.8% |  | 82 | 9.2% | 7.0% | 12.0% |  | 10 | 4.6% | 2.1% | 10.0% |  | 43 | 3.6% | 2.6% | 4.9% |  |
| **Marital status** |  |  |  |  | **0.002** |  |  |  |  | 0.513 |  |  |  |  | 0.100 |  |  |  |  | 0.373 |  |  |  |  | **0.003** |  |  |  |  | 0.771 |
| Unmarried | 707 | 6.4% | 5.7% | 7.2% |  | -- | -- | -- | -- |  | 124 | 9.8% | 7.6% | 12.5% |  | 330 | 9.6% | 8.3% | 11.2% |  | 67 | 3.4% | 2.6% | 4.5% |  | 186 | 3.1% | 2.7% | 3.7% |  |
| Currently married | 282 | 8.3% | 7.2% | 9.5% |  | 125 | 7.9% | 6.6% | 9.4% |  | 8 | 9.4% | 2.9% | 26.3% |  | 108 | 11.7% | 8.9% | 15.0% |  | 28 | 8.0% | 5.2% | 12.2% |  | 13 | 2.5% | 1.3% | 4.6% |  |
| Separated/Divorced/Widowed | 22 | 12.4% | 7.7% | 19.6% |  | 4 | 11.0% | 3.6% | 28.8% |  | 3 | 36.3% | 1.1% | 96.7% |  | 12 | 12.1% | 6.6% | 21.2% |  | 1 | 7.6% | Not estimable |  |  | 2 | 2.9% | 0.7% | 11.5% |  |
| **Children** |  |  |  |  | **0.063** |  |  |  |  | 0.180 |  |  |  |  | 0.981 |  |  |  |  | **0.044** |  |  |  |  | 0.039 |  |  |  |  | 0.670 |
| No children | 740 | 6.7% | 5.9% | 7.4% |  | 58 | 9.4% | 7.3% | 12.2% |  | 121 | 9.7% | 7.5% | 12.4% |  | 315 | 9.3% | 7.9% | 10.8% |  | 78 | 3.6% | 2.8% | 4.7% |  | 168 | 3.1% | 2.6% | 3.6% |  |
| Children | 208 | 8.1% | 6.8% | 9.5% |  | 53 | 7.3% | 5.5% | 9.5% |  | 11 | 9.6% | 4.7% | 18.4% |  | 104 | 12.3% | 9.6% | 15.5% |  | 12 | 7.5% | 3.9% | 13.8% |  | 28 | 3.4% | 2.2% | 5.1% |  |

Estimates are weighted to account for the DHS complex survey design. Country-specific estimates use DHS sampling weights, primary sampling units, and sampling strata. The pooled column represents a **country-normalised microdata estimate** (each country’s total weight is scaled to sum to 1) and should be interpreted as a **country-average pooled summary**. Prevalence is presented as % with design-based 95% confidence intervals. Unweighted sample sizes (n) are shown. Bangladesh contributes estimates for married females only.

Across the pooled sample, females reported thoughts of death or self-harm were three times as likely for females than for males (8.6% vs. 3.9%; p<0.001). This pattern was consistently observed across all countries where both sexes were surveyed and reached statistical significance in Lesotho, Mozambique, Nepal, and Zambia. Overall, urbanicity, educational attainment, and household wealth were not associated with thoughts of death or self-harm in the pooled analysis. A positive SES gradient was observed in Zambia; a negative gradient in Bangladesh.

In the pooled sample, adolescents who were currently married (8.5%) or separated/divorced/widowed (15.3%) had a much higher prevalence of thoughts of death or self-harm than unmarried adolescents (6.4%; p=0.007). Likewise, adolescents with children had a higher prevalence than those without children (10.4% vs. 6.3%; p=0.007). These associations were particularly evident in Mozambique and Nepal, whereas patterns varied across the remaining countries.

### Age gradients of probable depression and thoughts of death or self-harm

Figure 1 illustrates the age-specific prevalence of probable depression and thoughts of death or self-harm among adolescents aged 15–19 years by sex and country. Overall, age gradients were week. Depressive symptoms and thoughts of death or self-harm were generally more common among girls than boys at nearly all ages and in all countries where both sexes were surveyed.

**Figure 1.**
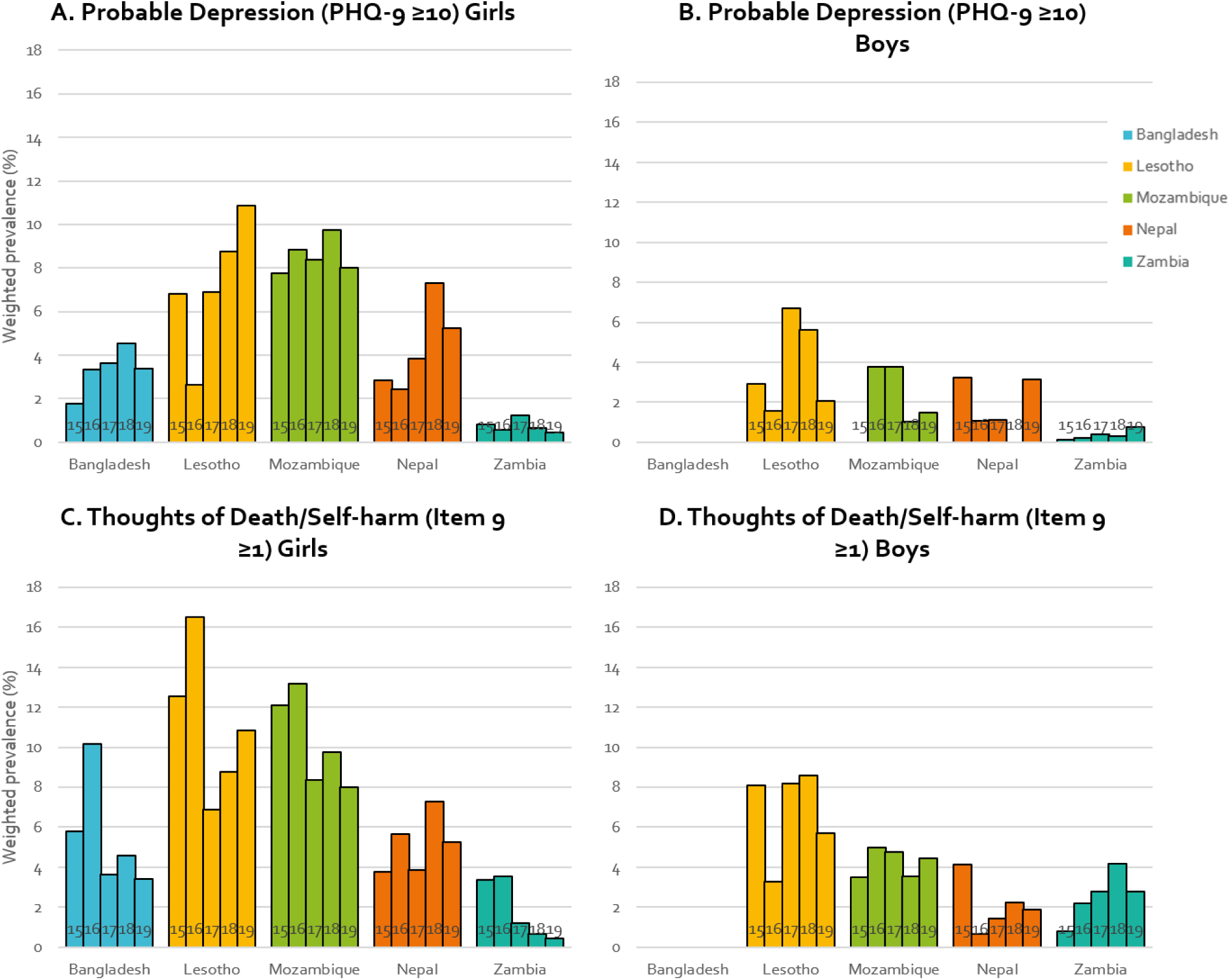
**Age-specific prevalence of probable depression (PHQ-9 ≥10) and thoughts of death or self-harm (PHQ-9 item 9 ≥1) across adolescents.** Bars show survey-weighted prevalence; vertical bars indicate 95% CIs. Panels are stratified by outcome and sex; each color represents a country (Bangladesh, Lesotho, Mozambique, Nepal, Zambia). Bangladesh contributes girls only because the mental health module was not administered to boys. Formal tests for age-related heterogeneity across ages 15–19 are reported in **Supplementary Table S2**.

Among girls, the prevalence of probable depression remained consistently highest in Mozambique across the adolescent years, while Lesotho showed a marked increase from age 16 onwards, reaching the highest prevalence at age 19. In contrast, prevalence remained comparatively low and stable in Zambia, and Bangladesh, while Nepal showed a modest increase in late adolescence. Among boys, age-related differences were less pronounced, with prevalence remaining low across most countries, except for a clear increase after 16 years old peaking at 17 in Lesotho and a transient peak in Mozambique during mid-adolescence.

Patterns for thoughts of death or self-harm were broadly similar but at substantially higher prevalence than probable depression as seen in Table 2. Among girls, Mozambique consistently showed the highest prevalence across all ages, whereas Lesotho demonstrated marked fluctuations, peaking at age 16 and 18. Bangladesh showed moderately higher prevalence in early years, while Nepal and Zambia remained stable across and comparatively lower. Among boys, prevalence was lower than among girls across all countries, although Lesotho and Mozambique again showed the highest estimates, with modest increases during late adolescence.

### Symptom profiles among adolescents by sex and country

Figure 2 compares PHQ-9 symptom endorsement patterns by country and sex among all adolescents and among those with probable depression. In Bangladesh, Nepal, and Zambia, fatigue or low energy (Item 4) was the most commonly endorsed symptom, whereas adolescents in Mozambique reported relatively higher endorsement of anhedonia (Item 1), depressed mood (Item 2), and sleep disturbance (Item 3). Girls in Lesotho showed a distinct profile characterized by comparatively greater endorsement of appetite changes (Item 5) and concentration difficulties (Item 7), with lower endorsement of fatigue than observed in Bangladesh and Nepal.

**Figure 2.**
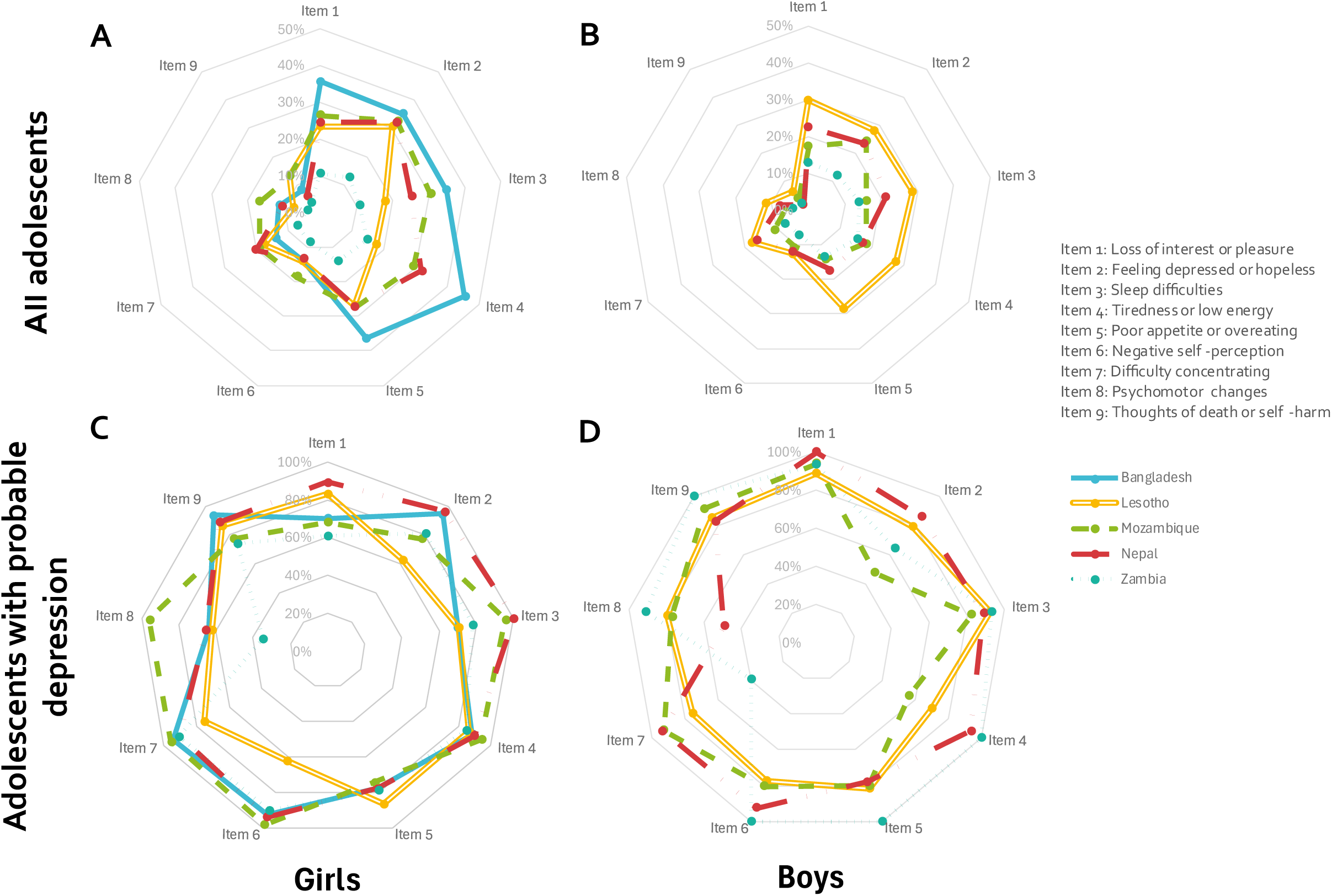
PHQ-9 symptom profiles among adolescents, overall and among those with probable depression, by sex and country. Spider plots display the weighted percentage of adolescents endorsing each PHQ-9 symptom at least “several days” in the past two weeks (item score ≥1). Panels show: (A) girls—overall sample, (B) boys—overall sample, (C) girls with probable depression (PHQ-9 ≥10), and (D) boys with probable depression (PHQ-9 ≥10). Each line represents a country (Bangladesh, Lesotho, Mozambique, Nepal, Zambia). Data callouts indicate the percentage of adolescents endorsing Item 9 (thoughts of self-harm or being better off dead) in each country and panel. PHQ-9 items are labeled numerically (Items 1–9); full item wording is provided in Supplementary Table S3. Estimates account for the DHS complex survey design and sampling weights. Bangladesh contributes girls only, as the mental health module was not administered to boys. Higher values indicate a greater share endorsing a given symptom, illustrating cross-country differences in symptom composition among adolescents overall and among those meeting the threshold for probable depression.

Among adolescents with probable depression (PHQ-9 ≥10), symptom profiles became substantially more homogeneous across countries, although notable item-level differences remained. Sleep disturbance (Item 3) was more frequently endorsed among girls with probable depression in Mozambique and Nepal than in Bangladesh or Lesotho, while the distinctive symptom profile observed in Lesotho persisted, with relatively greater endorsement of appetite and concentration symptoms. Among boys, symptom profiles were generally similar across countries, although greater between-country variability was observed than among girls because of smaller numbers. Across both sexes, endorsement of thoughts of death or self-harm (Item 9) remained high among adolescents with probable depression, exceeding 80% in most countries.

### Sociodemographic correlates of probable depression and thoughts of death or self-harm

Table 4 presents sex-stratified adjusted prevalence ratios for probable depression and thoughts of death or self-harm according to key sociodemographic characteristics.

**Table 4.**
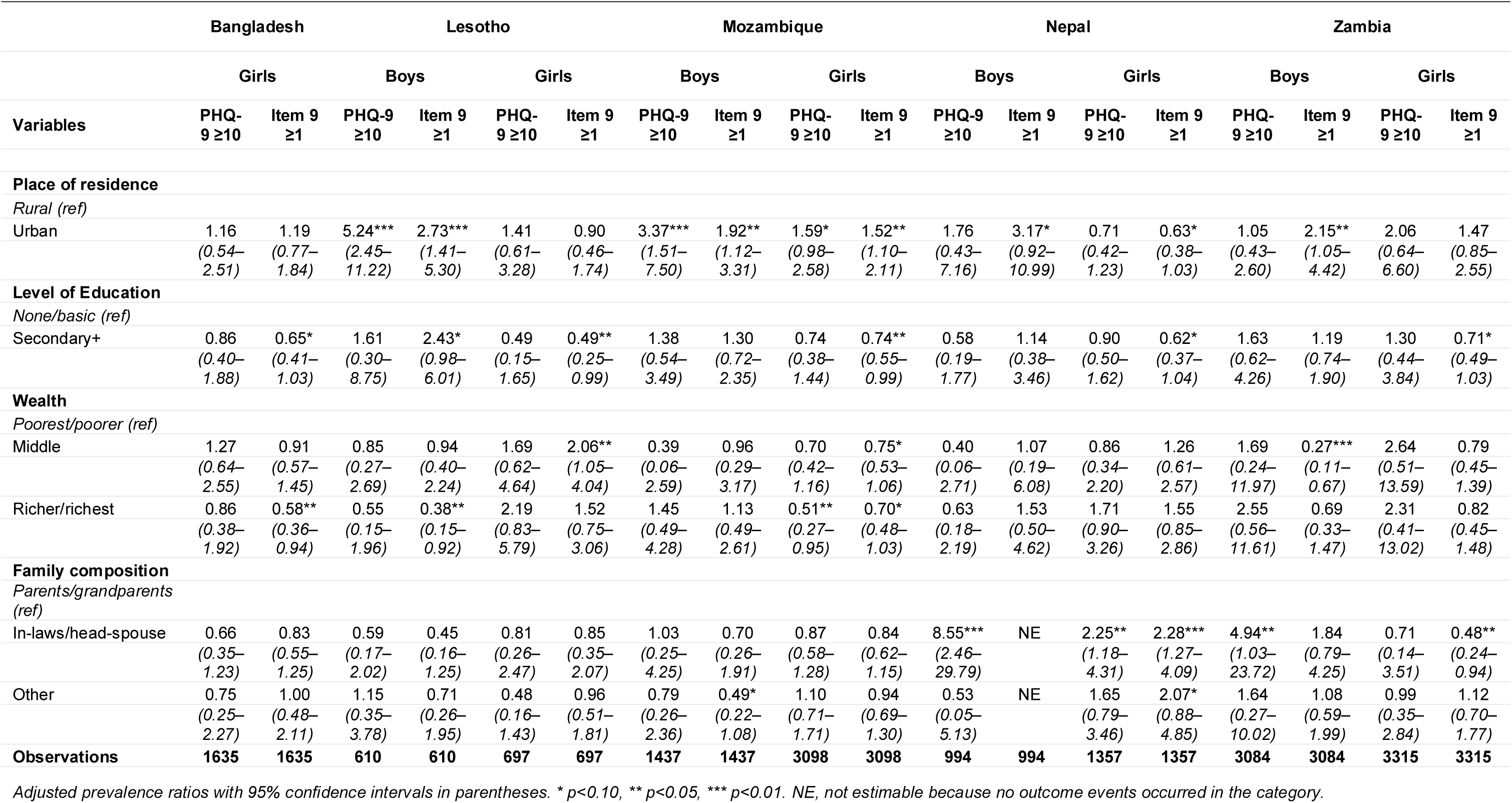
Adjusted prevalence ratios by country, sex, and outcome (15–19 years).

Urban residence was associated with a higher prevalence of mental health symptoms in several countries, particularly among boys. Urban boys in Lesotho had more than a fivefold higher prevalence of probable depression (aPR 5.24, 95% CI 2.45–11.22) and nearly threefold higher prevalence of thoughts of death or self-harm (aPR 2.73, 95% CI 1.41–5.30) than their rural counterparts. Similar positive associations were observed among boys in Mozambique and Nepal and among girls in Mozambique, whereas no association was observed in Bangladesh or Zambia.

Higher educational attainment showed predominantly protective associations, particularly for thoughts of death or self-harm. Adolescents with secondary or higher education had a lower prevalence of thoughts of death or self-harm in Bangladesh, Lesotho, Mozambique, Nepal, and Zambia, although statistically significant associations were not observed consistently across both sexes. In contrast, household wealth showed no clear gradient across countries or outcomes, with significant associations observed only in in Mozambique and Bangladesh with notably lower prevalence among wealthier adolescents.

Family composition emerged as the sociodemographic domain most strongly associated with mental health outcomes. Compared with adolescents living with parents or grandparents, those living with in-laws or as the head or spouse of their own household generally had higher prevalence of probable depression and thoughts of death or self-harm, particularly among girls in Nepal and boys in Zambia, although the magnitude and direction of associations differed across countries.

## Discussion

Using recent, nationally representative DHS data from 16,269 adolescents in five low- and middle-income ountries, this study provides, to our knowledge, the first cross-national comparison of probable depression nd thoughts of death or self-harm among adolescents aged 15–19 years using a common instrument in ationally representative household samples. Our analysis yielded three main findings. First, the prevalence f probable depression was relatively low in most countries, whereas thoughts of death or self-harm were more frequent. Second, girls consistently had a higher prevalence than boys where both sexes were urveyed. Third, neither single-year age patterns nor sociodemographic associations followed a consistent ross-country pattern, underscoring the importance of context-specific interpretation rather than assuming a niform adolescent mental-health profile across LMICs.

The relatively low prevalence of probable depression among adolescents was striking. Across the five urveys, fewer than one in 25 adolescents met the PHQ-9 threshold in the equally weighted pooled analysis, nd prevalence was below 1% in Zambia. These estimates are markedly lower than some pandemic-era meta-analytic estimates in which approximately one quarter of adolescents screened positive for depression (34) and seem only partially consistent with the global youth mental health crisis from WHO (3). Several actors likely explain this gap. Much of the currently available data is based on school-based, online or other onvenience samples using heterogeneous instruments, thresholds, and survey methods. By contrast, DHS urveys instead use household-based probability samples and include adolescents who are not enrolled in chool, providing a broader population perspective while limiting direct comparability with previous estimates.

A second challenge is the use of standardized cutoffs in the PHQ-9. While it is one of the most widely used nd extensively validated instruments for screening depressive symptoms in population-based mental health urveillance, it is a screening tool rather than a diagnostic interview with a score of ≥10 identifying probable epression rather than confirmed depressive disorder (35, 36). Although this threshold generally emonstrates good diagnostic accuracy, its sensitivity and specificity can vary by age, language, culture, and etting. Consequently, differences in translation, response styles, mental-health literacy, stigma, interviewer ffects, cultural adaptation, and willingness to disclose emotional distress may contribute to the present ariation between countries rather than reflecting true differences in the prevalence of depressive disorders 37, 38). Further, we do not know whether the cutoff of ≥10 is optimal in the populations studied because it as not been adequately validated in these adolescent populations. The estimates should consequently be nterpreted as standardized measures of PHQ-9-defined symptom burden rather than direct estimates of depressive disorder prevalence (39). In sensitivity analyses using the lower cutoff of ≥8, prevalence estimates ncreased across countries, although the overall pattern of cross-country and sex differences remained roadly similar (see Figure S5 in the supplementary material).

Documentation of validation for the specific PHQ-9 translations used in the DHS varied across countries. lthough country-specific validation studies exist for several settings, it is often unclear whether the DHS mployed the identical translated versions or whether comparable processes of linguistic and cultural daptation were followed. Formal assessment of measurement invariance across countries has also not been ndertaken. Accordingly, the marked differences observed between Zambia, Mozambique, and Lesotho might be interpreted as differences in PHQ-9-defined depressive symptom burden rather than definitive ankings of underlying depressive disorder. This consideration may be particularly important in settings with high burden of infectious diseases or undernutrition, where somatic symptoms such as fatigue or low energy may reflect physical illness in addition to depression (40, 41). Despite these limitations, the broadly similar ymptom profiles observed among adolescents meeting the PHQ-9 threshold across countries provide some eassurance that the instrument is capturing a comparable constellation of depressive symptoms in these diverse LMIC settings, although formal testing of measurement invariance is needed.

The finding that thoughts of death or self-harm were substantially more common than PHQ-9-defined robable depression in every country is particularly important. For context, WHO World Mental Health Survey ata from adults estimated 12-month major depressive episodes at 5.5% in high-income and 5.9% in low- to middle-income countries, compared with 12-month suicidal ideation at 2.0% and 2.1%, respectively (42, 43). lthough these diagnostic measures and recall periods are not directly comparable with our two-week PHQ-outcomes, the reverse ordering in our study is notable. This discrepancy may reflect a genuine pattern of dolescent psychological distress in which thoughts of death or self-harm occur even in the absence of levated depressive symptom scores. Although this relationship remains insufficiently understood in LMIC dolescent populations, the finding is consistent with evidence of substantial suicidal ideation and self-harm mong adolescents in several LMIC settings (15). It also highlights the importance of assessing suicide-elated thoughts independently of depression screening. It may also reflect differences in how depressive ymptoms and self-harm thoughts are understood or disclosed, or another indication of the PHQ-9 not working as well with this population.

HQ-9 Item 9 combines thoughts of being better off dead with thoughts of self-harm and cannot distinguish assive death wishes, non-suicidal self-injury, suicidal ideation, intent, or planning. The estimates should therefore not be interpreted as the prevalence of suicidal ideation or suicide risk. Nevertheless, endorsement dentifies adolescents experiencing potentially important distress and demonstrates that surveillance should ot rely on the PHQ-9 total-score threshold alone. Cultural understandings may also contribute to the ivergence: sadness, hopelessness, fatigue, or difficulty coping may be normalised or interpreted outside a mental-health framework. This may be particularly relevant among boys and young men, for whom norms mphasising strength, stoicism, and emotional control can discourage recognition or disclosure of depressive ymptoms (44, 45). Consequently, depressive symptoms captured by the PHQ-9 may be under-endorsed, while Item 9 may capture severe distress that is not necessarily recognised or expressed as part of a epressive syndrome. It is also important to highlight here that low prevalence of probable depression, as uggested by low PHQ-9 scores, does not rule out underlying concerns. Even modest percentages represent arge numbers of adolescents, particularly in settings with limited access to mental-health care.

The consistently higher prevalence observed among girls aligns with extensive evidence that sex differences n internalizing symptoms and depression emerge during adolescence and widen throughout the teenage ears (46). Potential explanations include differences in stress regulation and symptom reporting, gendered ocial norms, and greater exposure to interpersonal stressors and gender-based violence (47). Boys may be ess likely to acknowledge emotional distress or may express it through behaviours not captured by onventional depression screening (48). The observed differences may therefore reflect both a greater nderlying burden among girls and gendered patterns in the recognition and reporting of distress.

Among girls, early marriage and parenthood may capture major life transitions rather than demographic haracteristics alone. Their mental health implications are likely to depend on the circumstances in which hey occur. Early or unintended pregnancy, school discontinuation, economic dependence, limited partner or amily support, relationship conflict or violence, and substantial caregiving responsibilities may increase sychological distress (49–52). Conversely, marriage or motherhood may bring social recognition or family upport in some contexts (53). These associations should therefore be interpreted cautiously and in relation o the surrounding social and family environment. Some categories were uncommon and confidence intervals were wide. Moreover, the Bangladesh survey differed fundamentally from the others because its mental ealth module was administered only to ever-married adolescent girls, limiting direct comparison of marital tatus and childbearing.

The magnitude of the sex difference varied substantially across countries: it was smaller in Lesotho and ambia and approximately threefold for probable depression in Mozambique and Nepal. Context may help explain this heterogeneity, although the present data cannot test specific mechanisms. In Mozambique, ualitative evidence links early marriage and pregnancy with disrupted schooling, restricted opportunities, nd adverse emotional well-being among adolescent girls (54). In Lesotho, violence, orphanhood, economic ardship, and food insecurity may affect both sexes, while girls may face additional exposure to sexual or ntimate-partner violence, pregnancy, early marriage, and disrupted schooling (55, 56). In Nepal, greater omestic responsibilities (57) fewer educational opportunities and reduced mobility (58), and gender-based iolence (59), may increase girls’ distress. For example, a recent nationally representative survey consisting f 5,888 adolescents between the ages of 13-17 estimated that 3.9% had suicidal thoughts, a burden similar o our study, although it did not disaggregate this outcome by gender (60). Another nationwide school-based urvey, however, found that girls had 39% higher odds of suicidal ideation and 60% higher suicide attempt han boys (61). One possible contextual explanation for the smaller female-to-male differences observed in ambia and Lesotho is that countries may be at different stages of change in social norms surrounding masculinity and emotional expression. Traditional masculine norms emphasizing strength, self-reliance and motional restraint can reduce recognition and disclosure of psychological distress among boys (62, 63), whereas gender-transformative initiatives may gradually promote more flexible and positive constructions of masculinity. In Zambia, for example, the Coaching Boys Into Men programme has engaged adolescent boys, ncluding those aged 15–19 years, through mentorship and community dialogue to challenge harmful gender orms and promote positive masculinity; in Lesotho, MenEngage Lesotho and nationally institutionalised male-friendly services have similarly sought to engage men and boys, create safe spaces and challenge nequitable gender norms (64, 65). Such differences in the reach and maturity of gender-transformative rogramming could potentially influence boys’ willingness to recognise and report emotional distress, thereby ontributing to variation in observed sex differences between countries. These country-specific interpretations emain hypotheses, but they illustrate how gendered social environments and reporting practices may shape oth overall symptom burden and the size of observed sex differences.

We found little evidence of a common age gradient across adolescence. Apparent peaks at ages 17 or 18 were not sustained across adjacent ages and differed by country, sex, and outcome. Chronological age alone may therefore be a poor marker of mental health risk within the relatively narrow 15–19-year age range. A fe-course perspective where social transitions, rather than chronological age itself, are emphasized as markers of adolescent mental health risk may be more informative. This interpretation is consistent with vidence that adolescence is characterized by heterogeneous developmental trajectories, with the timing of educational, occupational, family, and caregiving transitions varying considerably across cultural and ocioeconomic contexts (66, 67).

Urban residence showed a more consistent association with thoughts of death or self-harm than with probable epression, although the pattern was not uniform across countries. Urban environments may expose dolescents to social fragmentation, academic and economic pressures, violence, or reduced social ohesion, while also affecting the recognition and reporting of distress(68, 69). For example, urban residence may facilitate greater recognition and reporting of emotional distress or reflect unmeasured differences in ocioeconomic and social environments between urban and rural settings (38).Greater exposure to digital nd social-media environments represents another possible pathway. Problematic social-media use and yberbullying have been associated with subsequent self-harm and suicidal behaviours among young people, lthough social media can also provide social support and protective messaging (70, 71). Because the DHS id not measure digital exposure or the proposed social mechanisms, these pathways remain hypotheses ather than explanations for the observed urban–rural differences.

The absence of a consistent association with household wealth should not be interpreted as evidence that material conditions are unimportant. Rather, it may reflect limitations of the DHS household asset index, which oes not capture dimensions of economic adversity particularly relevant to adolescents (72, 73), including ood insecurity, educational barriers, sudden household shocks, relative deprivation, or adolescents’ own ccess to financial resources (74–76). Material disadvantage may also influence mental health indirectly hrough pathways such as educational opportunities, household composition, family instability, or exposure o violence (77).

Overall, the sociodemographic correlates of adolescent mental health showed considerable heterogeneity cross countries, with few associations replicated consistently across settings. This finding suggests that road demographic characteristics alone are unlikely to identify adolescents at greatest risk across diverse MIC contexts. Instead, mental health appears to be shaped by context-specific social environments and life-ourse transitions, reinforcing the need for locally informed surveillance and prevention strategies rather than niversal risk profiles derived from pooled associations.

### Implications for surveillance and policy

These findings support continued investment in adolescent mental health in LMICs, particularly given the ubstantial prevalence of thoughts of death or self-harm and the limited availability of care. At the same time, hey highlight the need for greater precision in characterizing the global distribution of adolescent mental health problems. Estimates derived predominantly from high-income countries, school-based samples, or tudies conducted during the COVID-19 pandemic should not be generalized uncritically to adolescent opulations in LMICs. Conversely, relatively low prevalence estimates should not be interpreted as grounds or inaction, given the seriousness of suicidal thoughts and the large unmet need for mental health care in many LMICs.

A stronger surveillance strategy would combine repeated nationally representative household surveys with table instruments, inclusion of adolescents outside school, and embedded validation studies. Screening measures should be periodically calibrated against culturally adapted diagnostic interviews and measures of unctional impairment. Repeated use of the DHS mental health module could make an important contribution, articularly if future waves preserve comparable wording and administration procedures and include explicit valuation of language and measurement effects. Surveillance should also be linked to accessible responses or adolescents who report significant distress. This will require referral pathways connecting household urveys and community-based identification with adolescent-responsive primary care, schools, youth ervices, and community organisations. Existing youth-friendly platforms may provide practical entry points. n Mozambique, for example, the Serviços Amigos dos Adolescentes e Jovens (SAAJ) are integrated into rimary care and provide confidential counselling and health services for adolescents and young people (78). heir suitability for integrating mental health identification, brief support, and referral should be evaluated ather than assumed. Because many adolescents are outside school, responses should extend beyond ducational settings and involve coordinated action across health, education, social-protection, and community systems.

### Strengths and limitations

This study has several important strengths. It draws on large, nationally representative household surveys, roviding one of the first standardized descriptions of adolescent depressive symptoms across multiple MICs and including adolescents both in and out of school. The use of harmonized DHS survey methods and common measurement framework enhances cross-country comparability, while sex-stratified analyses, ge-specific estimates, and examination of both overall PHQ-9 scores and individual symptom profiles rovide a more nuanced characterization of adolescent mental health than prevalence estimates alone. inally, the use of a country-normalized pooled estimate prevented the largest national samples from isproportionately influencing the overall summary, allowing each country to contribute equally to cross-ountry comparisons.

Several limitations should be considered. The measures were self-reported and did not establish clinical iagnoses. Validation of the exact translated instruments varied across countries, and measurement nvariance was not tested. The item 9 outcome did not distinguish different forms or severity of suicidality. Country eligibility criteria differed, particularly in Bangladesh. Cross-sectional associations cannot establish emporality or causality, and sparse cells reduced precision in some subgroup analyses. Finally, the five ountries were included because suitable DHS data were available and should not be considered epresentative of LMICs as a whole; the pooled estimate is a descriptive benchmark for these surveys rather han a regional or global prevalence estimate.

### Conclusion

Our analysis of nationally representative data from five LMICs suggests that approximately 4% of dolescents had probable depression and 7% reported thoughts of death or self-harm. Both outcomes were more common among girls than boys, although the magnitude of these differences varied substantially cross countries. Age and sociodemographic associations were also heterogeneous, emphasizing the mportance of locally informed interpretation and responses. Further research should validate the PHQ-9 and its conventional cutoff across adolescent populations, improve the measurement of suicidal thoughts nd behaviours, and track changes in adolescent mental health using repeated population-based surveys.

## Supplemental Materials

- **Suppl Figure S1.** *Prevalence of probable depression (PHQ-9≥10) and thoughts of death or self-harm (PHQ-9 item 9≥1) by sex, pooled and by country. Bar chart*
- **Suppl Table S2.** Numeric prevalence tables for age gradients
- **Suppl Table S3.** Item endorsement tables
- **Suppl Figure S4.** Sex differences in individual PHQ-9 symptoms
- **Suppl Figure S5** cut-point sensitivity (PHQ-9 ≥8) Bar chart

## Data Availability

The minimal data set and accompanying code is available at Zenodo via https://zenodo.org/records/23726249

---

**Supplemental Figure S1.**
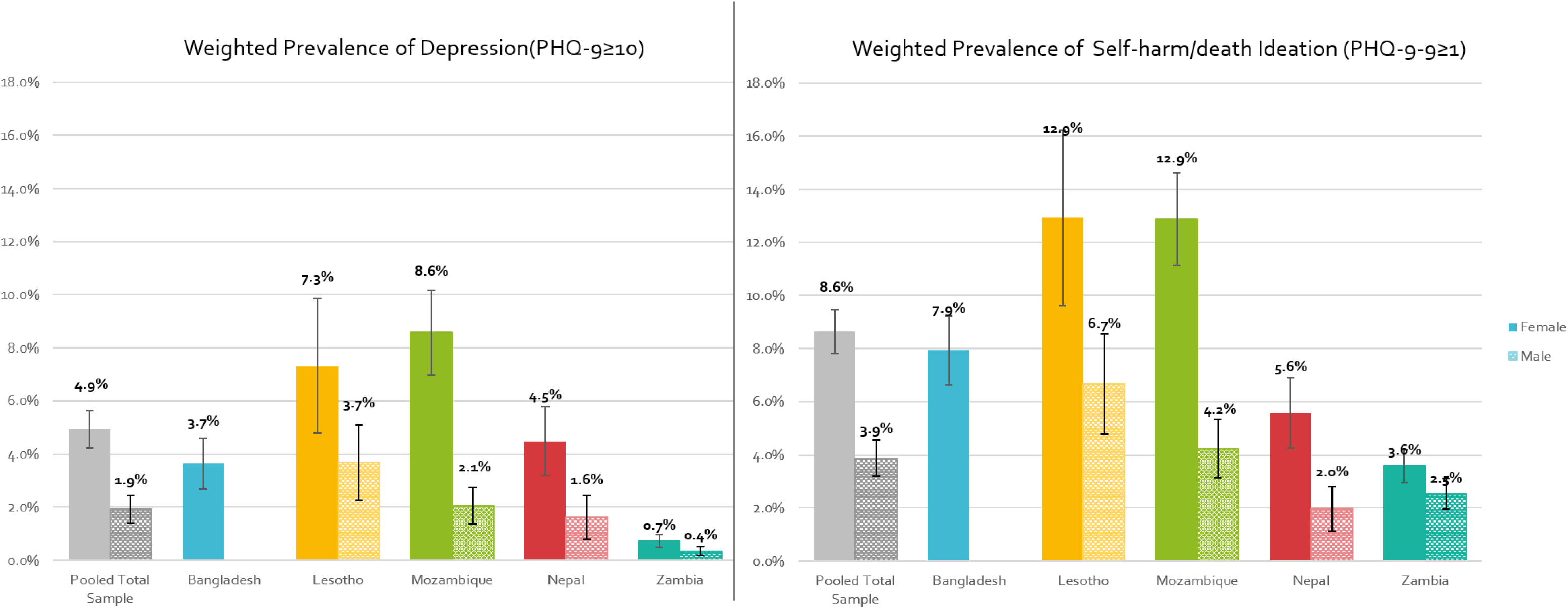
Prevalence of probable depression (PHQ-9≥10) and thoughts of death or self-harm (PHQ-9 item 9≥1) among adolescents aged 15–19 years by country and sex using country-specific design-based prevalence estimates. Pooled estimates are country-normalised microdata.

**Supplemental Figure S4.**
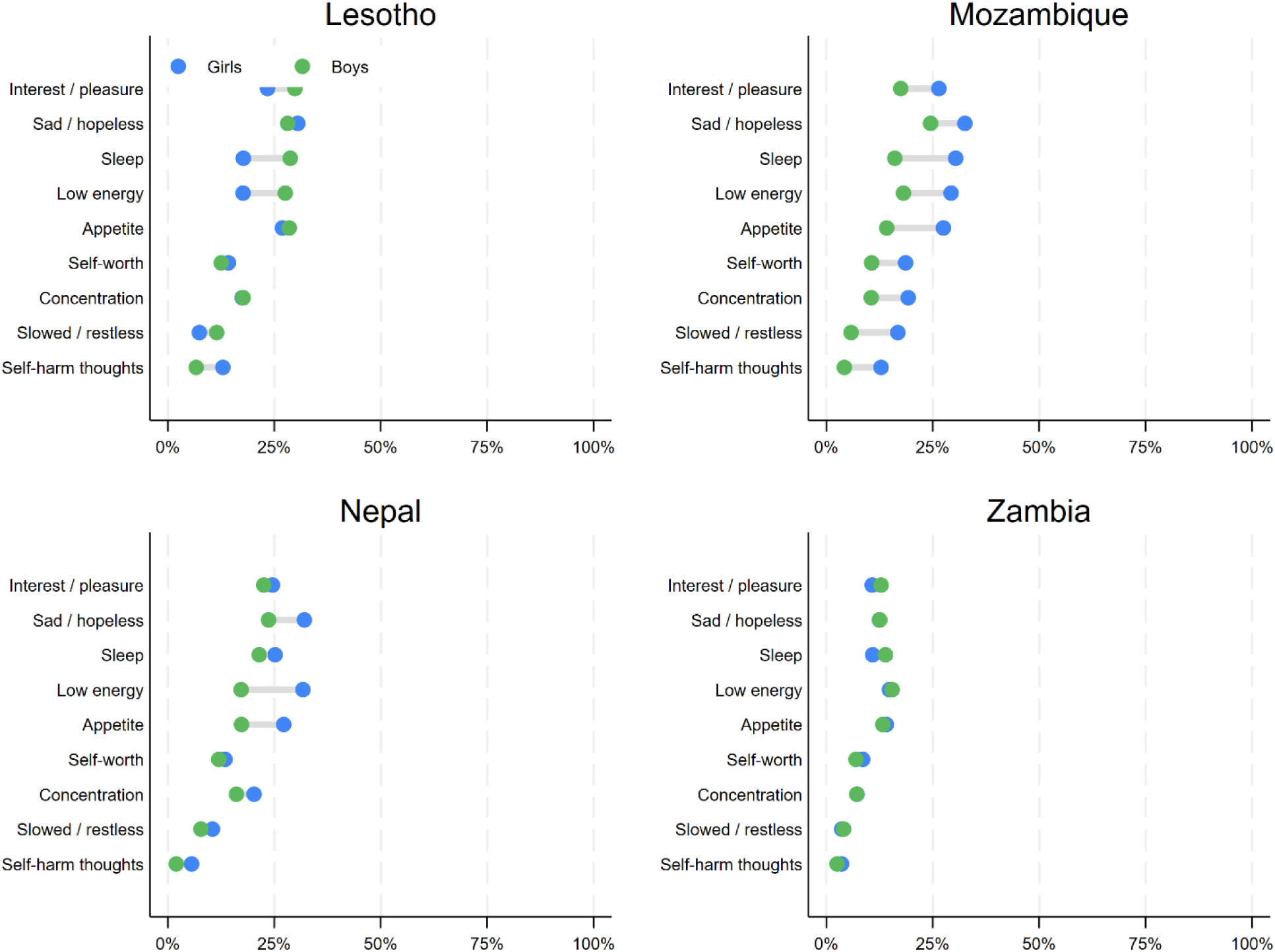
Sex differences in individual PHQ-9 symptoms among adolescents aged 15–19 years. Percentage-point differences in the weighted prevalence of each symptom between boys and girls are shown for all adolescents by country.

**Supplemental Table S2.** Prevalence of probable depression (PHQ-9 ≥10) and thoughts of death or self-harm (PHQ-9 item 9 ≥1) among adolescents aged 15–19 years. Heterogeneity tests by single-year age, sex, and country.

| Prevalence of probable depression (PHQ-9 ≥10) |  |  |  |  |  |  |  |  |  |  |  |  |  |  |  |
|---|---|---|---|---|---|---|---|---|---|---|---|---|---|---|---|
| Girls |  |  |  |  |  |  |  | Boys |  |  |  |  |  |  |  |
| Country | Age | n<br>(unweighted) | Prevalence | 95%CI |  | Age p-value* | Trend p-value** | Country | Age | n<br>(unweighted) | Prevalence | 95%CI |  | Age p-value* | Trend p-value** |
| Bangladesh | 15 | 3 | 1.8% | 0.5% | 6.2% | 0.653 | 0.448 |  |  |  |  |  |  |  |  |
| Bangladesh | 16 | 6 | 3.3% | 1.4% | 7.7% |  |  |  |  |  |  |  |  |  |  |
| Bangladesh | 17 | 8 | 3.6% | 1.8% | 7.2% |  |  |  |  |  |  |  |  |  |  |
| Bangladesh | 18 | 22 | 4.6% | 2.9% | 7.1% |  |  |  |  |  |  |  |  |  |  |
| Bangladesh | 19 | 17 | 3.4% | 1.9% | 5.9% |  |  |  |  |  |  |  |  |  |  |
| Lesotho | 15 | 5 | 6.8% | 2.1% | 19.7% | 0.342 | 0.229 | Lesotho | 15 | 3 | 4.6% | 1.3% | 15.2% | **0.041** | 0.813 |
| Lesotho | 16 | 3 | 2.6% | 0.6% | 10.8% |  |  | Lesotho | 16 | 3 | 1.6% | 0.5% | 4.9% |  |  |
| Lesotho | 17 | 8 | 6.9% | 2.7% | 16.4% |  |  | Lesotho | 17 | 8 | 6.7% | 3.1% | 13.9% |  |  |
| Lesotho | 18 | 15 | 8.8% | 4.7% | 15.8% |  |  | Lesotho | 18 | 11 | 5.6% | 3.5% | 8.9% |  |  |
| Lesotho | 19 | 12 | 10.8% | 5.7% | 19.6% |  |  | Lesotho | 19 | 5 | 2.1% | 0.9% | 4.8% |  |  |
| Mozambique | 15 | 31 | 8.3% | 5.5% | 12.5% | 0.934 | 0.933 | Mozambique | 15 | 0 | 0.0% | 0.0% | 0.0% | 0.111 | 0.718 |
| Mozambique | 16 | 34 | 8.7% | 5.9% | 12.7% |  |  | Mozambique | 16 | 10 | 3.8% | 2.0% | 7.1% |  |  |
| Mozambique | 17 | 33 | 8.3% | 5.6% | 12.2% |  |  | Mozambique | 17 | 11 | 3.8% | 2.0% | 7.0% |  |  |
| Mozambique | 18 | 51 | 9.7% | 6.9% | 13.5% |  |  | Mozambique | 18 | 4 | 1.0% | 0.4% | 2.8% |  |  |
| Mozambique | 19 | 36 | 8.0% | 5.7% | 11.1% |  |  | Mozambique | 19 | 5 | 1.9% | 0.8% | 4.6% |  |  |
| Nepal | 15 | 7 | 2.8% | 1.2% | 6.5% | 0.068 | **0.021** | Nepal | 15 | 5 | 3.2% | 1.2% | 8.2% | 0.439 | 0.855 |
| Nepal | 16 | 6 | 2.4% | 0.9% | 6.5% |  |  | Nepal | 16 | 2 | 1.1% | 0.3% | 4.3% |  |  |
| Nepal | 17 | 9 | 3.8% | 1.6% | 8.8% |  |  | Nepal | 17 | 2 | 1.1% | 0.3% | 4.6% |  |  |
| Nepal | 18 | 26 | 7.3% | 4.4% | 11.8% |  |  | Nepal | 18 | 0 | 0.0% | 0.0% | 0.0% |  |  |
| Nepal | 19 | 15 | 5.2% | 3.0% | 8.9% |  |  | Nepal | 19 | 4 | 3.1% | 0.7% | 13.3% |  |  |
| Prevalence of thoughts of death or self-harm (PHQ-9 item 9 ≥1) |  |  |  |  |  |  |  |  |  |  |  |  |  |  |  |
| Girls |  |  |  |  |  |  |  | Boys |  |  |  |  |  |  |  |
| Country | Age | n<br>(unweighted) | Prevalence | 95%CI |  | Age p-value* | Trend p-value** | Country | Age | n<br>(unweighted) | Prevalence | 95%CI |  | Age p-value* | Trend p-value** |
| Bangladesh | 15 | 9 | 5.8% | 2.9% | 11.3% | 0.608 | 0.593 |  |  |  |  |  |  |  |  |
| Bangladesh | 16 | 16 | 10.2% | 5.8% | 17.3% |  |  |  |  |  |  |  |  |  |  |
| Bangladesh | 17 | 24 | 9.5% | 6.3% | 13.9% |  |  |  |  |  |  |  |  |  |  |
| Bangladesh | 18 | 39 | 7.6% | 5.4% | 10.5% |  |  |  |  |  |  |  |  |  |  |
| Bangladesh | 19 | 41 | 7.3% | 5.2% | 10.0% |  |  |  |  |  |  |  |  |  |  |
| Lesotho | 15 | 13 | 12.5% | 6.3% | 23.4% | **0.013** | 0.936 | Lesotho | 15 | 10 | 10.5% | 5.4% | 19.6% | 0.256 | 0.579 |
| Lesotho | 16 | 15 | 16.5% | 8.4% | 30.0% |  |  | Lesotho | 16 | 7 | 3.6% | 1.6% | 7.8% |  |  |
| Lesotho | 17 | 11 | 5.4% | 3.0% | 9.5% |  |  | Lesotho | 17 | 10 | 8.2% | 4.2% | 15.3% |  |  |
| Lesotho | 18 | 30 | 19.2% | 12.0% | 29.2% |  |  | Lesotho | 18 | 14 | 8.6% | 5.1% | 14.0% |  |  |
| Lesotho | 19 | 19 | 10.0% | 5.6% | 17.1% |  |  | Lesotho | 19 | 11 | 5.7% | 2.9% | 10.9% |  |  |
| Mozambique | 15 | 65 | 12.2% | 9.1% | 16.2% | 0.633 | 0.723 | Mozambique | 15 | 8 | 3.5% | 1.5% | 8.1% | 0.883 | 0.862 |
| Mozambique | 16 | 76 | 13.5% | 10.1% | 17.7% |  |  | Mozambique | 16 | 15 | 5.1% | 3.1% | 8.4% |  |  |
| Mozambique | 17 | 69 | 11.6% | 8.7% | 15.2% |  |  | Mozambique | 17 | 18 | 4.8% | 2.9% | 7.7% |  |  |
| Mozambique | 18 | 107 | 14.5% | 11.5% | 18.2% |  |  | Mozambique | 18 | 11 | 3.5% | 1.7% | 7.3% |  |  |
| Mozambique | 19 | 75 | 12.6% | 9.8% | 16.0% |  |  | Mozambique | 19 | 12 | 4.8% | 2.9% | 8.0% |  |  |
| Nepal | 15 | 9 | 3.8% | 1.8% | 7.7% | 0.691 | 0.364 | Nepal | 15 | 8 | 4.1% | 1.8% | 9.3% | 0.275 | 0.620 |
| Nepal | 16 | 17 | 5.7% | 3.2% | 9.9% |  |  | Nepal | 16 | 2 | 0.7% | 0.1% | 3.1% |  |  |
| Nepal | 17 | 15 | 6.0% | 3.4% | 10.5% |  |  | Nepal | 17 | 3 | 1.4% | 0.4% | 4.8% |  |  |
| Nepal | 18 | 24 | 6.9% | 4.2% | 11.4% |  |  | Nepal | 18 | 2 | 2.2% | 0.5% | 8.8% |  |  |
| Nepal | 19 | 14 | 5.0% | 2.9% | 8.4% |  |  | Nepal | 19 | 2 | 1.9% | 0.5% | 7.1% |  |  |

*Note*\*Age p-value is the omnibus test for any differences in prevalence across single-year ages 15–19 within each country and sex stratum (design-based Wald test for the categorical age indicator).
\*\*Trend p-value is the test for a linear trend with age (design-based Wald test treating age 15–19 as an ordered/continuous term). Both p-values come from survey-weighted models that account for the DHS complex sampling design (weights, clustering, and stratification)

**Supplementary Table S3.**
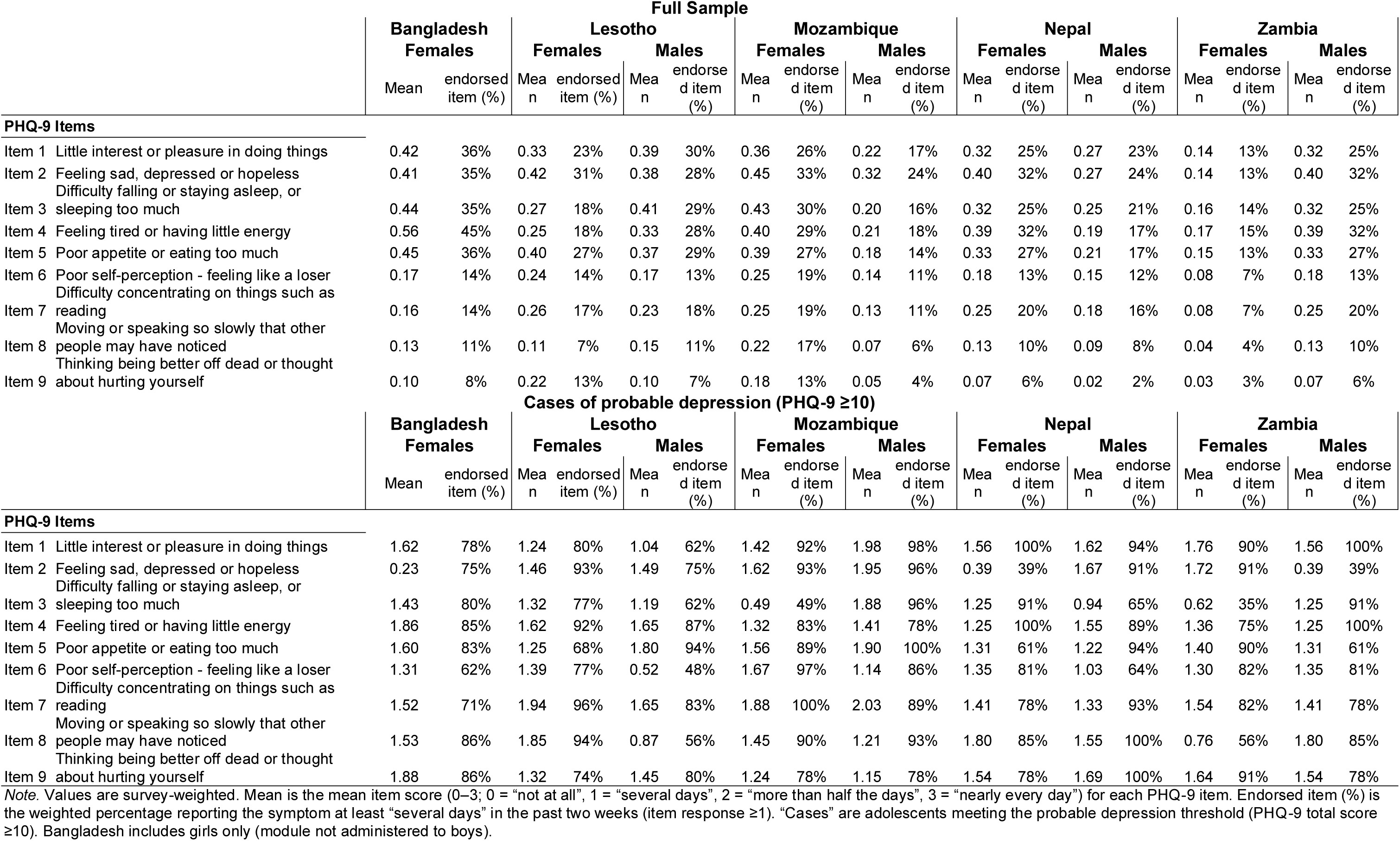
PHQ-9 item response profiles among adolescents aged 15–19 years, by sex and country (full sample and cases of probable depression).

| Full Sample |  |  |  |  |  |  |  |  |  |  |  |  |  |  |  |  |  |  |  |
|---|---|---|---|---|---|---|---|---|---|---|---|---|---|---|---|---|---|---|---|
|  |  | **Bangladesh** |  | **Lesotho** |  |  |  | **Mozambique** |  |  |  | **Nepal** |  |  |  | **Zambia** |  |  |  |
|  |  | **Females** |  | **Females** |  | **Males** |  | **Females** |  | **Males** |  | **Females** |  | **Males** |  | **Females** |  | **Males** |  |
|  |  | Mean | endorsed item (%) | Mean | endorsed item (%) | Mean | endorsed item (%) | Mean | endorsed item (%) | Mean | endorsed item (%) | Mean | endorsed item (%) | Mean | endorsed item (%) | Mean | endorsed item (%) | Mean | endorsed item (%) |
| **PHQ-9 Items** |  |  |  |  |  |  |  |  |  |  |  |  |  |  |  |  |  |  |  |
| Item 1 | Little interest or pleasure in doing things | 0.42 | 36% | 0.33 | 23% | 0.39 | 30% | 0.36 | 26% | 0.22 | 17% | 0.32 | 25% | 0.27 | 23% | 0.14 | 13% | 0.32 | 25% |
| Item 2 | Feeling sad, depressed or hopeless | 0.41 | 35% | 0.42 | 31% | 0.38 | 28% | 0.45 | 33% | 0.32 | 24% | 0.40 | 32% | 0.27 | 24% | 0.14 | 13% | 0.40 | 32% |
| Item 3 | Difficulty falling or staying asleep, or sleeping too much | 0.44 | 35% | 0.27 | 18% | 0.41 | 29% | 0.43 | 30% | 0.20 | 16% | 0.32 | 25% | 0.25 | 21% | 0.16 | 14% | 0.32 | 25% |
| Item 4 | Feeling tired or having little energy | 0.56 | 45% | 0.25 | 18% | 0.33 | 28% | 0.40 | 29% | 0.21 | 18% | 0.39 | 32% | 0.19 | 17% | 0.17 | 15% | 0.39 | 32% |
| Item 5 | Poor appetite or eating too much | 0.45 | 36% | 0.40 | 27% | 0.37 | 29% | 0.39 | 27% | 0.18 | 14% | 0.33 | 27% | 0.21 | 17% | 0.15 | 13% | 0.33 | 27% |
| Item 6 | Poor self-perception - feeling like a loser | 0.17 | 14% | 0.24 | 14% | 0.17 | 13% | 0.25 | 19% | 0.14 | 11% | 0.18 | 13% | 0.15 | 12% | 0.08 | 7% | 0.18 | 13% |
| Item 7 | Difficulty concentrating on things such as reading | 0.16 | 14% | 0.26 | 17% | 0.23 | 18% | 0.25 | 19% | 0.13 | 11% | 0.25 | 20% | 0.18 | 16% | 0.08 | 7% | 0.25 | 20% |
| Item 8 | Moving or speaking so slowly that other people may have noticed | 0.13 | 11% | 0.11 | 7% | 0.15 | 11% | 0.22 | 17% | 0.07 | 6% | 0.13 | 10% | 0.09 | 8% | 0.04 | 4% | 0.13 | 10% |
| Item 9 | Thinking being better off dead or thought about hurting yourself | 0.10 | 8% | 0.22 | 13% | 0.10 | 7% | 0.18 | 13% | 0.05 | 4% | 0.07 | 6% | 0.02 | 2% | 0.03 | 3% | 0.07 | 6% |
| **Cases of probable depression (PHQ-9 ≥10)** |  |  |  |  |  |  |  |  |  |  |  |  |  |  |  |  |  |  |  |
|  |  | **Bangladesh** |  | **Lesotho** |  |  |  | **Mozambique** |  |  |  | **Nepal** |  |  |  | **Zambia** |  |  |  |
|  |  | **Females** |  | **Females** |  | **Males** |  | **Females** |  | **Males** |  | **Females** |  | **Males** |  | **Females** |  | **Males** |  |
|  |  | Mean | endorsed item (%) | Mean | endorsed item (%) | Mean | endorsed item (%) | Mean | endorsed item (%) | Mean | endorsed item (%) | Mean | endorsed item (%) | Mean | endorsed item (%) | Mean | endorsed item (%) | Mean | endorsed item (%) |
| **PHQ-9 Items** |  |  |  |  |  |  |  |  |  |  |  |  |  |  |  |  |  |  |  |
| Item 1 | Little interest or pleasure in doing things | 1.62 | 78% | 1.24 | 80% | 1.04 | 62% | 1.42 | 92% | 1.98 | 98% | 1.56 | 100% | 1.62 | 94% | 1.76 | 90% | 1.56 | 100% |
| Item 2 | Feeling sad, depressed or hopeless | 0.23 | 75% | 1.46 | 93% | 1.49 | 75% | 1.62 | 93% | 1.95 | 96% | 0.39 | 39% | 1.67 | 91% | 1.72 | 91% | 0.39 | 39% |
| Item 3 | Difficulty falling or staying asleep, or sleeping too much | 1.43 | 80% | 1.32 | 77% | 1.19 | 62% | 0.49 | 49% | 1.88 | 96% | 1.25 | 91% | 0.94 | 65% | 0.62 | 35% | 1.25 | 91% |
| Item 4 | Feeling tired or having little energy | 1.86 | 85% | 1.62 | 92% | 1.65 | 87% | 1.32 | 83% | 1.41 | 78% | 1.25 | 100% | 1.55 | 89% | 1.36 | 75% | 1.25 | 100% |
| Item 5 | Poor appetite or eating too much | 1.60 | 83% | 1.25 | 68% | 1.80 | 94% | 1.56 | 89% | 1.90 | 100% | 1.31 | 61% | 1.22 | 94% | 1.40 | 90% | 1.31 | 61% |
| Item 6 | Poor self-perception - feeling like a loser | 1.31 | 62% | 1.39 | 77% | 0.52 | 48% | 1.67 | 97% | 1.14 | 86% | 1.35 | 81% | 1.03 | 64% | 1.30 | 82% | 1.35 | 81% |
| Item 7 | Difficulty concentrating on things such as reading | 1.52 | 71% | 1.94 | 96% | 1.65 | 83% | 1.88 | 100% | 2.03 | 89% | 1.41 | 78% | 1.33 | 93% | 1.54 | 82% | 1.41 | 78% |
| Item 8 | Moving or speaking so slowly that other people may have noticed | 1.53 | 86% | 1.85 | 94% | 0.87 | 56% | 1.45 | 90% | 1.21 | 93% | 1.80 | 85% | 1.55 | 100% | 0.76 | 56% | 1.80 | 85% |
| Item 9 | Thinking being better off dead or thought about hurting yourself | 1.88 | 86% | 1.32 | 74% | 1.45 | 80% | 1.24 | 78% | 1.15 | 78% | 1.54 | 78% | 1.69 | 100% | 1.64 | 91% | 1.54 | 78% |

*Note.* Values are survey-weighted. Mean is the mean item score (0–3; 0 = “not at all”, 1 = “several days”, 2 = “more than half the days”, 3 = “nearly every day”) for each PHQ-9 item. Endorsed item (%) is the weighted percentage reporting the symptom at least “several days” in the past two weeks (item response ≥1). “Cases” are adolescents meeting the probable depression threshold (PHQ-9 total score ≥10). Bangladesh includes girls only (module not administered to boys).

**Supplemental Figure S5.**
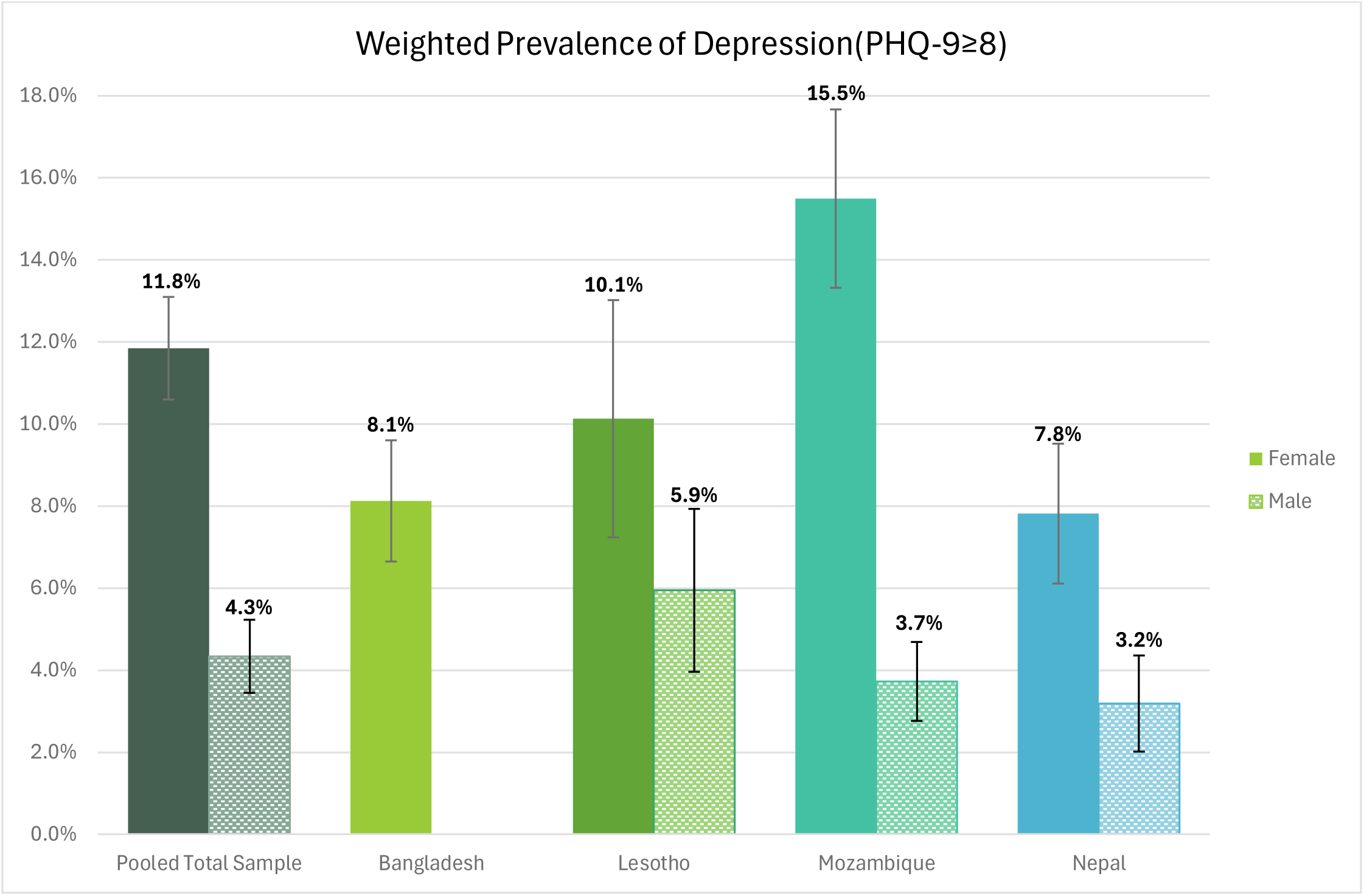
Cut-point sensitivity (PHQ-9 ≥8). Survey-weighted prevalence of depressive symptoms among adolescents by sex and country. Solid bars represent females and patterned bars represent males. Using this alternate threshold (PHQ≥8), prevalence was higher among females relative to males in the pooled and in individual countries (Lesotho, Mozambique and Nepal). Males were excluded in Bangladesh. Error bars indicate 95% confidence intervals.

